# Urinary metabolic biomarkers of diet quality in European children are associated with metabolic health

**DOI:** 10.1101/2021.09.09.21263335

**Authors:** Nikolaos Stratakis, Alexandros P. Siskos, Eleni Papadopoulou, Anh N. Nguyen, Yinqi Zhao, Katerina Margetaki, Chung-Ho E. Lau, Muireann Coen, Lea Maitre, Silvia Fernández-Barrés, Lydiane Agier, Sandra Andrusaityte, Xavier Basagaña, Anne Lise Brantsaeter, Maribel Casas, Serena Fossati, Regina Grazuleviciene, Barbara Heude, Rosemary R C McEachan, Helle Margrete Meltzer, Christopher Millett, Fernanda Rauber, Oliver Robinson, Theano Roumeliotaki, Eva Borràs, Eduard Sabidó, Jose Urquiza, Marina Vafeiadi, Paolo Vineis, Trudy Voortman, John Wright, David V. Conti, Martine Vrijheid, Hector C. Keun, Leda Chatzi

## Abstract

Urinary metabolic profiling is a promising powerful tool to reflect dietary intake and can help understand metabolic alterations in response to diet quality. Here, we used ^1^H-NMR spectroscopy in a multi-country study in European children (1147 children from 6 different cohorts) and identified a common panel of 4 urinary metabolites (hippurate, *N*-methylnicotinic acid, urea and sucrose) that was predictive of Mediterranean diet adherence (KIDMED) and ultra-processed food (UPF) consumption and also had higher capacity in discriminating children’s diet quality than that of established sociodemographic determinants. Further, we showed that the identified metabolite panel also reflected the associations of these diet quality indicators with C-peptide, a stable and accurate marker of insulin resistance and future risk of metabolic disease. This methodology enables objective assessment of dietary patterns in European child populations, complementary to traditional questionary methods, and can be used in future studies to evaluate diet quality. Moreover, this knowledge can provide mechanistic evidence of common biological pathways that characterize healthy and unhealthy dietary patterns, and diet-related molecular alterations that could associate to metabolic disease.

## Introduction

Dietary habits are considered a key element for the prevention of chronic non-communicable diseases.^1^ In 2017, 22% of all deaths among adults, were attributed to dietary risks, with type 2 diabetes being among the top causes of diet-related deaths.^2^ However, it is notoriously difficult to measure diet accurately in large population studies and the absence of accurate dietary assessment methods is hampering the evidence linking diet and disease.^3, 4^ There is a need for novel approaches to better elucidate diet-related metabolic alterations and their association with disease risk.

Metabolomics is the systematic study of small-molecule metabolites in a biological system and has recently emerged as a powerful top-down approach providing a comprehensive phenotype of biological status. Urinary metabolic phenotypes carry rich information on environmental, lifestyle and nutritional exposures, physiological and metabolic status, and disease risks on an individual and population level.^5–7^ Urine specimens have high concentrations of food-derived metabolites and studies have shown that urinary metabolic profiles could provide an objective measure of dietary intake.^8^

Previous research identifying diet-related metabolic profiles, has largely focused on selected food groups including fruits, vegetables, meat and seafood,^5, 9–12^ while dietary patterns and food processing are far less studied.^6, 7, 13, 14^ Ultra-processed foods (UPFs), which are industrial formulations undergoing a series of physical and chemical processes and typically lack intact healthy food components and include various additives, can result to cumulative intake of salt, added sugars, and fats.^15^ UPF consumption has been increasing worldwide^16–19^ and, to our knowledge, no previous report exists on its metabolic signature. In contrast with the study of overall diets, including the UPF diet, exploring intakes of single foods, which is what is traditionally done in nutrition research, might be failing to provide a realistic image of dietary metabolic footprint. Moreover, most previous studies have focused on adults, and little is known about body’s metabolic response to diet during childhood. This is important as age is a major source of variation in metabolite profiling, with large differences being observed between children and adults.^20^

Alterations in metabolite profiling could provide a mechanistic link between diet and disease development as early as in childhood. Alterations in the levels of several metabolic biomarkers and pathways have been associated with insulin resistance in childhood including, branched-chain and aromatic amino acid metabolism, urea cycle, glucose and carbohydrate metabolism.^21–23^ In addition, dietary patterns characterized by high consumption of UPFs, such soft drinks, sweet and savoury snacks, have been related to higher risk of insulin resistance in children.^24, 25^ All previous studies have included measurement of serum insulin levels as marker of insulin resistance. An alternative marker is C-peptide, a protein that is co-secreted with insulin on an equimolar basis from pancreatic beta cells and has been shown to strongly predict metabolic disease progression.^26^ C-peptide has a longer half-life than insulin and is recognized as a stable and accurate marker of endogenous insulin secretion, even in non-fasting conditions.^27–29^ Previous studies in children have shown that higher carbohydrate intake is related to higher C-peptide concentrations.^30, 31^ However, there is little evidence on the metabolic signatures underlying the association of diet with C-peptide levels in children. We conducted a multi-country study in European children within the Human Early-Life Exposome (HELIX) project,^32^ aiming *1*) to identify urinary metabolites associated with Mediterranean diet adherence and UPF consumption, and *2)* to determine the extent to which these metabolites were associated with C-peptide, used as an early marker of metabolic health.

## Methods

### Study population

This study is embedded within the Human Early-Life Exposome (HELIX) project,^32^ a collaborative project across six established and ongoing longitudinal population-based birth cohort studies in Europe: Born in Bradford (BiB, United Kingdom),^33^ Étude des Déterminants pré et postnatals du développement et de la santé de l’Enfant (EDEN, France),^34^ Kaunas Cohort (KANC, Lithuania),^35^ INfancia y Medio Ambiente (INMA, Spain),^36^ Norwegian Mother, Father and Child Cohort Study (MoBa, Norway),^37^ and RHEA (RHEA, Greece).^38^ Participating cohorts covered singleton deliveries from 2003 to 2008. As part of HELIX, a sub-cohort of 1301 children (approximately 200 children in each cohort) were followed in 2014 to 2015 for a clinical examination, a computer-assisted interview with the parents, and the collection of biological samples. Data collection was standardized across cohorts and performed by trained staff. A full description of the HELIX follow-up methods and study population are provided by Maitre et al.^32^

Our study population consisted of 1147 children with available information on dietary intake, plasma C-peptide levels, and metabolomic biomarkers in urine gathered during the HELIX follow-up at a mean age of 7.9 years (range: 5.4-12.0 years) (Supplementary Figure 1). All participating families provided written informed consent. Approval for the HELIX project was obtained from the local ethical committees at each site. Additionally, the current study was approved by the University of Southern California Institutional Review Board.

### Dietary assessment

Information about the children’s habitual diet was collected via a semi quantitative food-frequency questionnaire (FFQ) covering the child’s habitual diet, which was filled in by the parent attending the examination appointment. The FFQ, covering the past year, was developed by the HELIX research group, translated and applied to all cohorts.^32^ It included 43 questions of intake of food items, which were aggregated in 16 main food groups: meat & meat products, fish & seafood, sweets, beverages, potatoes, vegetables, dairy products, fruits, bread & cereal, sweet bakery products, added fats, eggs, nuts, salty snacks, pulses and dressings. It also included 15 specific questions to examining the degree of adherence to Mediterranean diet. Diet quality was assessed using two different approaches, i) by assessing the degree of adherence to a Mediterranean diet based on the KIDMED index (Mediterranean Diet Quality Index for children and adolescents)^39^ and ii) by assessing the proportion of UPF in the overall diet.^15^

For the KIDMED index,^39^ items positively associated with the Mediterranean diet pattern (11 items) were assigned a value of +1, while those negatively associated with the Mediterranean diet pattern (4 items) were assigned a value of −1 (Supplementary Table 1). The scores for all 15 items were summed, resulting in a total KIDMED score ranging from −4 to 11, with higher scores reflecting greater adherence to a Mediterranean diet. We categorized the score into three groups: low (<1); moderate (1-4); and high (>4).

For ultra-processed food (UPF) intake, we identified foods and drinks as “ultra-processed” by using the NOVA classification, a food classification system based on the nature, extent, and purpose of industrial food processing.^15^ We identified the following “ultra-processed” foods: cookies, pastries, sugar-sweetened, low-sugar and artificially-sweetened beverages, cold meat cuts; ham, dairy desserts, sugar-sweetened and other breakfast cereals, crispbread and rusks; chocolate, sweets, margarine, dressings and salty snacks. For some food items, our FFQ did not provide enough information on food processing to determine if a specific item belongs to one processing category or another. We discussed the classification of each food item with a team of nutritionists and used a conservative approach, such that the lower level of processing was chosen-for instance, we made the assumption that fries are homemade from fresh potatoes, and therefore, they were not classified as UPF. For each child, we calculated the daily proportion of all UPF in the total diet as the ratio between the sum of daily servings of UPF to the total daily sum of all food and drink servings. More details on the categorization of foods according to the NOVA classification are presented in Supplementary Table 2.

### Urine metabolite profiling

Two urine samples, representing last night-time and first morning voids, were collected on the evening and morning before the clinical examination, kept in a fridge and transported in a temperature-controlled environment, and aliquoted and frozen within 3 h of arrival at the clinics. They were subsequently pooled to generate a more representative sample of the last 24 h for metabolomic analysis ^9^.

Urinary metabolic profiles were acquired using ^1^H NMR spectroscopy according to ^9^. In brief one-dimensional 600 MHz ^1^H NMR spectra of urine samples from each cohort were acquired on the same Bruker Avance III spectrometer operating at 14.1 Tesla within a period of 1 month. The spectrometer was equipped with a Bruker SampleJet system, and a 5-mm broad-band inverse configuration probe maintained at 300K. Prior to analysis, cohort samples were randomised. Deuterated 3-(trimethylsilyl)-[2,2,3,3-d_4_]-propionic acid sodium salt (TSP) was used as internal reference. Aliquots of the study pooled quality control (QC) sample were used to monitor analytical performance throughout the run and were analysed at an interval of every 23 samples (i.e. 4 QC samples per well plate). The ^1^H NMR spectra were acquired using a standard one-dimensional solvent suppression pulse sequence. 44 metabolites were identified and quantified as described in ^9^. The urinary NMR showed excellent analytical performance, the mean coefficient of variation across the 44 NMR detected urinary metabolites was 11%. For the statistical analysis we have used creatinine-normalized metabolite concentrations (μmol/mmol of creatinine).

### Plasma C-peptide

Blood was collected at the end of the clinical examination during the HELIX follow-up visit. The median postprandial interval (time between last meal and blood collection) was 3.3 h (IQR: 2.8–4.0).

For each cohort, concentration of C-peptide was assessed in child plasma at the CRG/UPF Proteomics Unit (Barcelona, Spain) using the xMAP and Luminex System multiplex platform according to the manufacturer’s protocol. Blood samples were randomized and blocked by cohort prior to measurement to ensure a representation of each cohort in each plate (batch). For protein quantification, an 8-point calibration curve per plate was performed with protein standards provided in the Luminex kit and following the procedures described in the standard procedures described by the vendor. Commercial heat inactivated, sterile-filtered plasma from human male AB plasma (Sigma Cat #. H3667) was used as constant controls to control for intra-and inter-plate variability. Four control samples were added per plate. Raw intensities obtained with the xMAP and Luminex system for each sample were converted to pg/mL using the calculated standard curves of each plate and accounting for the dilutions that were made prior measurement. The coefficient of variation for C-peptide was 16%. The LOD was determined and the lower and upper quantification limits (LOQ1 and LOQ2, respectively) were obtained from the calibration curves. C-peptide concentrations were log2-transformed to achieve normal distribution. Plate batch effect was corrected by subtracting for each individual and each protein the difference between the overall protein average minus the plate specific protein average. Finally, values below LOQ1 and above LOQ2 were imputed using a truncated normal distribution using the truncdist R package.

### Covariates

Adjustment factors were selected *a priori* based on literature^40–42^ and included: maternal age (in years), maternal education level (low, middle, high), maternal pre-pregnancy BMI (in kg/m^2^), family affluence score (cohort-specific definition of low, middle, high), child sex, child age (in years), child BMI (in kg/m^2^), child sedentary behavior (minutes/day of time spent watching TV, playing computer games or other sedentary games), child ethnicity (White European, Asian, other), and postprandial interval (in hours). We also included a cohort indicator as a fixed effect in the models, as this, in the context of an observational study, is expected to control for cohort effects.^43^ We imputed missing values for covariates (ranging from 0 to 4%) using the method of chained equations with the R package *mice*. Details about the imputation process in HELIX, diagnostics, and comparison between imputed and complete-case values have been reported in detail elsewhere.^44^

### Statistical analysis

As a first step in our analysis, we conducted a metabolome-wide association study (MWAS) to assess the associations of urinary metabolites with diet quality. Creatinine-normalized metabolite concentrations (μmol/mmol of creatinine) were log_10_ transformed prior to statistical analyses to improve model fit. We fitted separate multivariable regression models for each metabolite with the KIDMED score or UPF intake (expressed as per 5% change of total daily food intake). To account for multiple hypothesis testing, we applied the Benjamin–Hochberg false discovery rate (FDR) correction; an FDR-corrected *P* value <0.05 denoted statistical significance. For metabolites identified to be associated with the diet quality indicators, we assessed between-cohort heterogeneity with the I^2^ statistic and χ2 test from Cochran’s Q.

To examine the ability of the identified metabolite panels in discriminating children with low *vs.* high KIDMED scores (<1 *vs* >4) and low *vs.* high UPF intakes (Quartile 1: <18% *vs* Quartile 4: ≥29%), we plotted receiver operating characteristic (ROC) curves and estimated area under the ROC curve (AUC) values, indicative of the discriminative performance of the metabolite models, based on ten-fold cross validations. We also repeated the ROC analysis for a set of established sociodemographic factors (maternal education level, maternal pre-pregnancy BMI, family affluence score, child sedentary behavior, ethnicity, age, and sex) linked to childhood diet quality both previously^40–42^ and in our study population (all P<0.05) and compared the discriminative performance of this sociodemographic set with that of the metabolites.

Next, we examined the association of the KIDMED score and of UPF intake (as independent variables) with plasma C-peptide concentration (as dependent variable) using multivariable linear regression models. No departures from linearity in the associations of these diet quality indicators with C-peptide concentration were observed both visually and statistically (P for linearity > 0.42) using generalized additive models. We examined the KIDMED score both as continuous (per score unit increase) and in categories of low (score<1, reference), moderate (score=1-4), and high (score>4). Likewise, UPF intake was assessed both as continuous (per 5% change of total daily food intake) and in quartiles (Q1: <18%, reference; Q2: 18%-<23%; Q3: 23%-<29%; and Q4: ≥29% of total daily food intake). We also included a product term between KIDMED and UPF intake in the regression analysis to assess their interaction; to simplify interpretation of this model, we categorized the KIDMED score as low/moderate *vs* high and UPF intake based on the median population intake (<23% *vs.* ≥23%). We conducted two sets of sensitivity analyses to assess the robustness of the results. First, we calculated cohort-specific effect estimates and assessed heterogeneity with the I^2^ statistic and χ2 test from Cochran’s Q. Second, we examined potential effect modification by child sex and by child weight status (IOTF-defined normal weight vs overweight/obese) on C-peptide by testing the multiplicative interaction term between the potential effect modifier and each diet quality measure. Finally, we fitted regression models with the metabolites found to be associated with diet quality indicators, and C-peptide in order to assess whether diet-related metabolites are related to β-cell function.

We performed analyses with both complete (missingness <4% in each covariate) and imputed data. Results were similar across raw and imputed data analyses, and hence, we present those using the imputed covariate data. For easier interpretation of effect estimates for the log transformed C-peptide and metabolite values, we back-transformed regression coefficients and present results as percent change (% change =(back-transformed[beta] – 1) × 100).

Analyses were conducted using STATA version 14.2 (StataCorp LLC, TX) and R software version 3.5.3. Linear regression analyses were performed in STATA with the command “*regress” and using “mi estimate”* to account for the imputed covariate data.^45^ ROC analysis was performed in R with the caret package.^46^ Visualizations of the results were carried out using the ggplot2 package in R.^47^

## Results

### Study population

Among participating children (n=1147), 626 (54.6%) were boys and 1028 (89.6%) were white (Table 1). The mean (SD) age at assessment was 7.9 (1.6) years. Median (IQR) C-peptide concentration was 1.26 (0.03, 1.95) ng/mL.

**Table 1.** Characteristics of the study population.

|  |  |
| --- | --- |
| **Cohort of inclusion, n (%)** |  |
| BiB, UK | 189 (16.5) |
| EDEN, France | 149 (13) |
| INMA, Spain | 202 (17.6) |
| KANC, Lithuania | 194 (16.9) |
| MoBa, Norway | 221 (19.3) |
| RHEA, Greece | 192 (16.7) |
| **Maternal characteristics** |  |
| Maternal age, mean (SD), years | 30.7 (4.9) |
| Missing, n (%) | 13 (1.1) |
| Pre-pregnancy BMI, mean (SD), kg/m<sup>2</sup> | 25 (5) |
| Missing, n (%) | 21 (1.8) |
| Maternal educational level, n (%) |  |
| Low | 157 (13.7) |
| Medium | 391 (34.1) |
| High | 562 (49) |
| Missing, n (%) | 37 (3.2) |
| **Child characteristics** |  |
| Age at assessment, mean (SD), years | 7.9 (1.6) |
| Sex, n (%) |  |
| Male | 626 (54.6) |
| Female | 521 (45.4) |
| Ethnicity, n (%) |  |
| White European | 1028 (89.6) |
| Asian | 92 (8) |
| Other | 27 (2.4) |
| Family Affluence Score, n (%) |  |
| Low | 126 (11) |
| Medium | 448 (39.1) |
| High | 569 (49.6) |
| Missing, n (%) | 4 (0.4) |
| BMI, mean (SD), kg/m<sup>2</sup> | 16.9 (2.6) |
| Normal weight, n (%)<sup>1</sup> | 906 (79) |
| Overweight/obese, n (%)<sup>1</sup> | 237 (20.7) |
| Missing, n (%) | 4 (0.4) |
| KIDMED score, mean (SD) | 2.8 (1.7) |
| Low (<1), n (%) | 104 (9.1) |
| Medium (1-4), n (%) | 848 (73.9) |
| High (>4), n (%) | 195 (17) |
| Ultra-processed food intake, mean (SD), % of daily food intake | 24.2 (8.7) |

Abbreviations: BiB, Born in Bradford cohort; EDEN, the Étude des Déterminants pré et postnatals du développement et de la santé de l'Enfant study; INMA, Infancia y Medio Ambiente cohort; KANC, Kaunas Cohort; KIDMED, Mediterranean Diet Quality Index for children and adolescents; MoBa, Norwegian Mother, Father and Child Cohort Study; RHEA, Rhea Mother Child Cohort study.
<sup>1</sup> Categories of normal weight and overweight/obese were derived using the International Obesity Taskforce criteria.<sup>72</sup>

Seventeen percent of children (n=195) had a high KIDMED score (>4), indicative of high adherence to the Mediterranean diet. Mean (SD) UPF intake in the overall study population was 24.2 (8.7) % of total daily food intake. KIDMED and UPF intake were negatively correlated (Spearman r = −0.44); children with high and low (<1) KIDMED score had a mean (SD) UPF intake of 18.8 (6.6) % and 33.4 (9.9) %, respectively. The two diet quality scores were associated with most recorded food intakes in opposite directions (Supplementary Tables 3 and 4). Daily fruit and vegetable consumption, weekly fish consumption and not skipping breakfast were the major dietary habits differentiating children with low and high KIDMED score (Supplementary Table 1). Intakes of pastries and bakery products, dairy desserts, margarine and dressings were the major determinants of UPF intake (Supplementary Figure 2). Children with the highest KIDMED scores were mostly from Norway and Spain (Supplementary Figure 3), while those with the highest UPF intake were mostly from Lithuania and the UK (Supplementary Figure 4).

### Diet quality and urinary metabolome

Figure 1 and Supplementary Tables 5 and 6 show the association of diet quality indicators with urinary metabolite levels in childhood. KIDMED and UPF intake exhibited an opposite pattern of association for most of the metabolites (30 out of 43). After controlling for false discovery rate (FDR-corrected P value<0.05), we found that a panel of 4 metabolites related to both diet quality indicators. Specifically, a higher KIDMED score was associated with higher levels of hippurate, *N*-methylnicotinic acid, and urea and with lower levels of sucrose; UPF intake exhibited opposite associations with these 4 metabolites. A higher KIDMED score was also associated with higher acetate and pantothenic acid concentrations, while tyrosine and valine concentrations were inversely associated with UPF intake. There was no evidence of significant between-cohort heterogeneity in the associations between the diet quality indicators and these metabolites (I^2^ <30%; p for heterogeneity >0.2). ROC curve analyses showed that the combination of 4 metabolites associated with both diet quality indicators performed better than individual metabolites in discriminating children with high and low KIDMED scores and UPF intake (Figure 2). The discriminative ability of this metabolite panel was not improved with the addition of urinary metabolites specifically linked to each diet quality indicator and was equal or even greater to that of established sociodemographic factors previously linked to diet quality in childhood. The regression formulas (scores) for predicting children’s diet quality indicators based on the urinary metabolites are given in Supplementary Table 7.

**Figure 1.**
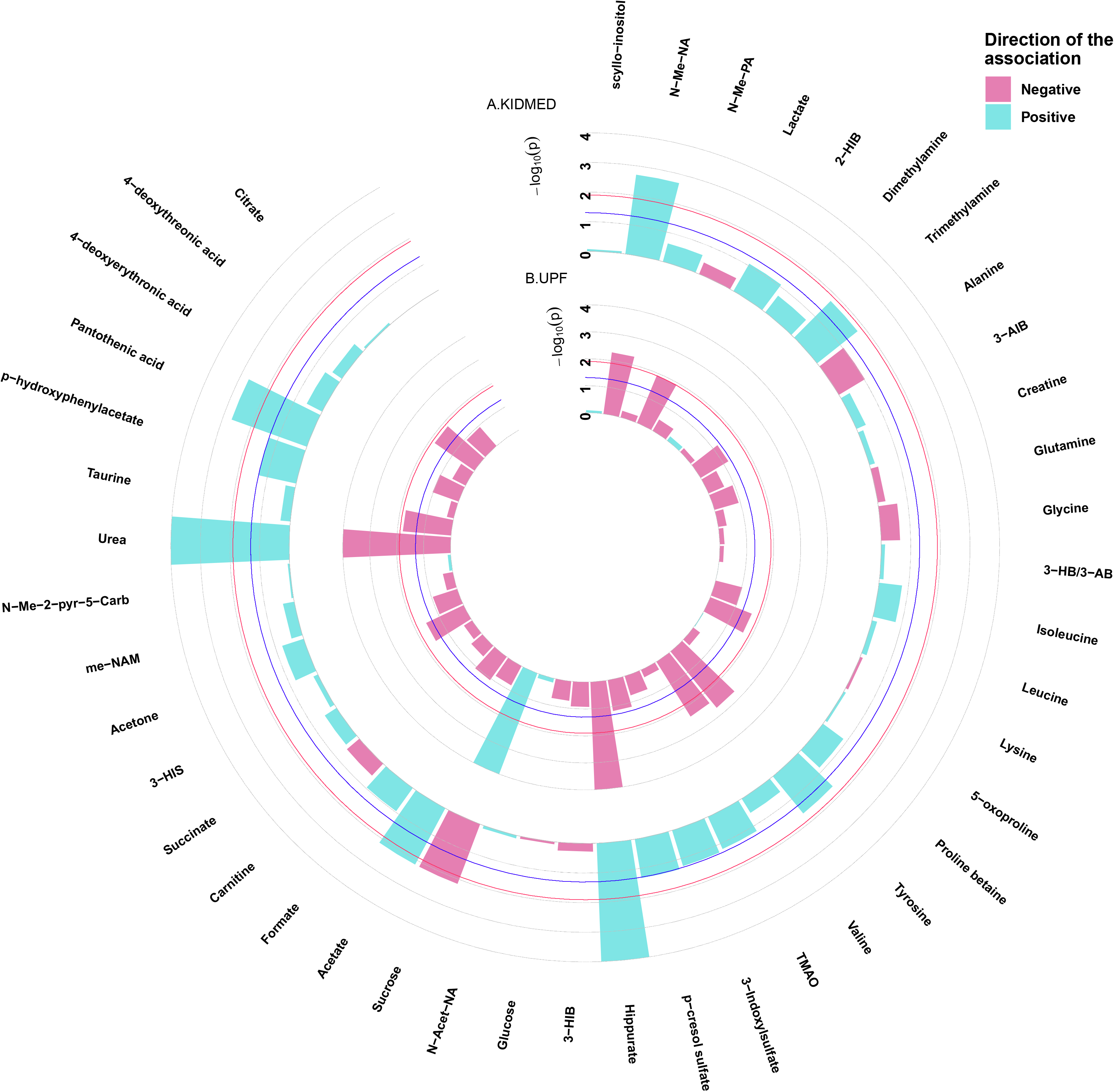
Adjusted associations of diet quality indicators with urinary metabolites in childhood. Linear regression models were adjusted for maternal age, maternal education level, maternal pre-pregnancy BMI, family affluence status, child sex, child age, child BMI, child sedentary behavior, child ethnicity, and a cohort indicator. The blue line represents a P-value of 0.05. The red line represents an FDR-adjusted P-value of 0.05. 2-HIB, 2-hydroxyisobutyrate; 3-AIB, 3-aminoisobutyrate; 3-HB/3-AB, 3-hydroxybutyrate/3-aminoisobutyrate; 3-HIB, 3-hydroxyisobutyrate; 3-HIS, 3-hydroxyisovalerate; me-NAM, N1-methyl-nicotinamide; *N*-Acet-NA, *N*-acetyl neuraminic acid; *N*-Me-2-pyr-5-Carb, *N*-methyl-2-pyridone-5-carboxamide; *N*-me-NA, *N*-methylnicotinic acid; *N*-me-PA, *N*-methylpicolinic acid; TMAO, Trimethylamine *N*-oxide.

**Figure 2.**
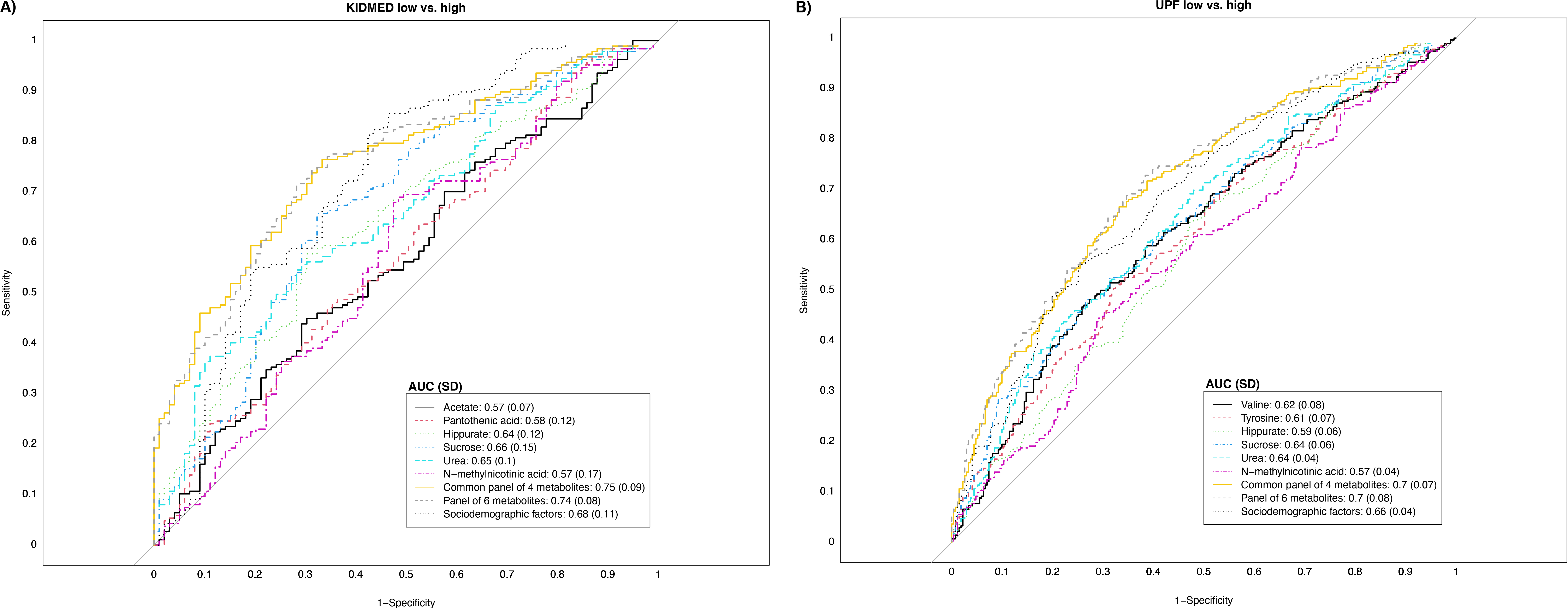
Receiver operating characteristic (ROC) curves reflecting the ability of urinary metabolites of interest in discriminating adherence to diet quality in childhood. Panel A illustrates the ability of urinary metabolites of interest in discriminating high adherence to the Mediterranean diet (KIDMED > 4) from low adherence (KIDMED <1). Panel B illustrates the ability of urinary metabolites of interest in discriminating high ultra-processed food consumption (UPF ≥29% of total intake) from low consumption (UPF <18% of total intake). The area under the receiver operating characteristic curve (AUC) values is displayed in the legend. The common panel of 4 metabolites includes the metabolites associated with both diet quality indicators (hippurate, sucrose, urea and *N*-methylnicotinid acid). The panel of 6 metabolites includes the metabolites associated with each diet quality indicator (common panel of 4 plus acetate and pantothenic acid for KIDMED, and plus valine and tyrosine for UPF). The panel of sociodemographic factors includes maternal education level, maternal pre-pregnancy BMI, family affluence score, child sedentary behavior, ethnicity, age, and sex.

### Diet quality and C-peptide levels

Table 2 presents the associations of diet quality indicators with C-peptide concentration in childhood. We found that a higher KIDMED score was associated with lower C-peptide. Specifically, compared to a low KIDMED score (<1), children with a moderate score (1-4) had a 28% lower C-peptide concentration (percent change: −27.7, 95% CI: −49.6 to 3.9) and those with a high score (>4) had a 39% lower C-peptide concentration (percent change: −39.0, 95% CI: −60.6 to −5.7) (P-trend =0.03). An opposite association was observed for UPF intake. Compared to children at the lowest quartile of UPF intake (<18% of total daily food intake), children at the second quartile (18-<23% of total daily food intake) had a 24% higher C-peptide concentration (percent change: 24.3, 95% CI: −6.4 to 65.2), those at the third quartile (23-<29% of total daily food intake) had a 39% higher concentration (percent change: 38.5, 95% CI: 3.8 to 84.9), and those at the fourth quartile (≥29% of total daily food intake) had a 46% higher concentration (percent change: 46.0, 95% CI: 8.1 to 97.3) (P-trend=0.01). There was no evidence of interaction between the diet quality indicators (Supplementary Table 8). When we examined the associations separately in each cohort, we found no significant between-cohort heterogeneity (I^2^<18%, p for heterogeneity >0.29) (Supplementary Figures 5 and 6). We also found no evidence that the observed associations differed by the sex of the children or their weight status (Supplementary Table 9).

**Table 2.** Adjusted associations of diet quality with C-peptide levels in childhood^1^.

|  | **C-peptide** |
| --- | --- |
|  | **Percent change (95% CI)** |
| KIDMED score (per unit increase) | -8.1 (-13.7, -2.2) |
| Low (<1) | *Ref.* |
| Moderate (1-4) | -27.7 (-49.6, 3.9) |
| High (>4) | -39.0 (-60.6, -5.7) |
| P-trend | 0.03 |
| UPF intake (per 5% increase of total intake) | 9.3 (2.8, 16.2) |
| Q1 (<18% of total intake) | *Ref.* |
| Q2 (18- <23% of total intake) | 24.3 (-6.4, 65.2) |
| Q3 (23-<29% of total intake) | 38.5 (3.8, 84.9) |
| Q4 (≥29% of total intake) | 46.0 (8.1, 97.3) |
| P-trend | 0.01 |

Abbreviations: KIDMED, Mediterranean Diet Quality Index for children and adolescents; UPF, ultra-processed food
<sup>1</sup> Effect estimates represent percent changes in log-2 transformed C-peptide levels and their 95% CIs derived from linear regression models adjusted for maternal age, maternal education level, maternal pre-pregnancy BMI, family affluence status, child sex, child age, child BMI, child sedentary behavior, child ethnicity, postprandial interval, and a cohort indicator.

### Diet-related urinary metabolites and C-peptide levels

When we examined the associations with the metabolite scores for each diet quality indicator, we found that the scores for KIDMED were associated with lower C-peptide, while opposite associations were observed with the metabolite scores for UPF (Supplementary Table 10). In the pairwise associations between the individual urinary metabolites linked to KIDMED or UPF intake and C-peptide, we found that higher levels of sucrose were associated with higher C-peptide levels

## Discussion

This is the first study of European children that identified a panel of urinary metabolites associated with diet quality, as assessed by a Mediterranean diet adherence score (KIDMED) and ultra-processed food (UPF) intake. These metabolites include food constituents (or their metabolic products), vitamin related compounds and metabolites related to amino acid, protein and carbohydrate metabolism. For both diet quality indices, KIDMED score and UPF intake, there was a common panel of 4 metabolites exhibiting an opposite pattern of association: higher levels of hippurate, *N*-methylnicotinic acid and urea, and lower levels of sucrose were concomitant with a higher KIDMED score and lower UPF intake. Moreover, these urinary metabolites that reflected these diet quality indicators, were also associated with C-peptide levels, a well-known marker of-cell function and insulin resistance.^26^

Previous studies assessing the metabolic response of the body to diet have largely focused on specific foods groups,^6^ yet humans eat a combination of foods that may have additive or interactive effects on human physiology. In our study, we assessed two diet quality indices to better describe the complexity of overall diet. The Mediterranean diet is characterized by high intake of fruits, vegetables, legumes, nuts, and whole grain products, fish and low intakes of red meat and sweets, and it has long been appraised for its cardiometabolic benefits, even in children.^48^ Moreover, we assessed UPF intake to characterize the intake of industrial processed and prepared food items, with altered food structure, nutritional content, and taste^49^. UPF intake assessment diverts from the traditional strategy of studying nutrients, foods or dietary patterns to identify the link between diet with health and disease, and it reflects the cumulative intake of artificial substances (i.e. flavorings, colorings, emulsifiers, and other additives) and processing by-products. Effects of UPF consumption are likely to be the result of synergistic effects of many food ingredient compound and characteristics, and high consumption of UPF has been proposed to lead to metabolic dysregulation.^50^

For both KIDMED and UPF, there was a common panel of 4 urinary metabolites (hippurate, *N*-methylnicotinic acid, urea, sucrose), exhibiting an opposite pattern of association. In addition, KIDMED was positively associated with pantothenic acid and acetate, whereas UPF was negatively associated with two amino acids, valine and tyrosine. These metabolic signatures demonstrate a core common metabolic biomarker panel reflecting habitual dietary intake, but also show that the two diet quality indices and their metabolic signatures act complementary and can highlight different aspects of human metabolism and physiology. Our finding are consistent with previous metabolomics studies conducted in adults.^13, 51, 52^ Four dietary interventions were developped^13^ with similar energy content and within the WHO healthy eating guidelines, but with varying macro-and micronutrient intake. The urine metabolic profile for the diet most concordant with the guidelines, characterized by high intakes of dietary fiber (through fruits, vegetables and whole grain cereal products) and low intakes of fat, sugar and salt showed systematic differences for a total of 28 metabolites, including increased levels of acetate, hippurate, *N*-methylnicotinic acid and urea, compared to the diet that diverted from the healthy guidelines.^13^ The PREDIMED study also used NMR to define urinary biomarkers associated with a high adherence to a Mediterranean diet pattern in adults and the proposed biomarkers also included higher levels of urea.^52^ To our knowledge, our study is the first to show the urinary metabolic footprint of total ultra-processed food intake, and also the first to show that urine NMR-derived scores of diet quality are reflective of a key biomarker of metabolic health, c-peptide, in healthy children. The ability to use a rapid, non-invasive biofluid screen to measure objective biomarkers of diet quality in children opens new avenues for exploring the significance of nutritional patterns to healthy development early in life.

Among the diet-related urinary metabolites, sucrose individually reflected the associations observed between diet quality and C-peptide.^53^ Elevated levels of added sugars could lead to elevated glucose load in the human body, which in turn can lead to an increase in glycemic response and, thus, C-peptide production in healthy populations. To the best of our knowledge, there is only one previous study from Mexico examining the relation of diet to C-peptide in children.^54^ Similar to our findings, this study showed that adherence to a prudent dietary pattern characterized by food groups commonly consumed in Mediterranean diet (vegetables, fruit, fish, and legumes) was associated with low C-peptide levels in boys. Regarding other markers of glucose regulation, our findings are in line with previous studies in children and adults reporting that low adherence to a healthy dietary pattern or high consumption of specific ultra-processed foods (sugar-sweetened beverages, and ultra-processed meat) were associated with impaired glucose regulation and insulin resistance.^55–60^ Overall, our findings are consistent with the public health concerns raised by the World Health Organization (WHO), relating poor overall quality with high intake level of sugars and added sugars, and with poor metabolic health and a high risk of several non-communicable diseases.^53, 61–63^

Regarding the other urinary metabolites reflecting diet quality, hippurate is a normal component of urine, a metabolic product of phenolic compounds which are present in various dietary sources. It is also a biomarker of fruit/vegetable intake, as it has been confirmed in previous studies in healthy children, adolescents, ^9, 64^ and adults.^65^ *N*-methylnicotinic acid (trigonelline) is a product of the metabolism of niacin (vitamin B3) and a biomarker of various dietary sources like legumes^66^ and fruits.^9^ Urea is the principal end product of amino acids and protein catabolism and a potential marker of protein intake from food.^51^

In addition, adherence to the Mediterranean diet was also positively associated with urinary levels of pantothenic acid and acetate. Pantothenic acid (vitamin B5, necessary to form coenzyme-A) is present in many foods, and we have previously reported a positive association between consumption of dairy products and urinary pantothenic acid, in the same study population.^9^ Acetate has also multiple dietary sources, it is produced by acetate producing bacteria in foodstuff and urinary acetate is also modulated by human gut microbial metabolism. We have previously reported a positive association between potato consumption and urinary acetate levels, in the same study population.^9^ Both compounds have a central role in human biochemistry and the metabolism and synthesis of carbohydrates, proteins, and fats.

Moreover, UPF intake was negatively associated with two amino acids, valine and tyrosine. Tyrosine is regarded as a conditionally essential amino acid in adults and essential in children. Foods high in dietary tyrosine include dairy, meat, eggs, beans, nuts, grains. Tyrosine is a precursor for neurotransmitters and hormones, increases dopamine availability which in turn could enhance cognitive performance.^67^ Valine is an essential branch chain amino acid critical to energy homeostasis, protein and muscle metabolism.^68, 69^ Neither of these metabolites have been linked with habitual dietary intake before.

### Strengths and limitations

The main strengths of the study are the multicentric design which included children from 6 countries spanning north to south in Europe, the use of standardized data collection and biomarker measurement protocols across cohorts, and the fairly large sample size with biomarker data. Identified panels of urinary metabolites had similar or higher capacity in discriminating children’s diet quality to that of established sociodemographic determinants. We chose ^1^H NMR spectroscopy for urinary analysis as this is an inherently high reproducible, high throughput technique suitable for the identification and quantification of urine metabolites, which are typically of high concentrations, without complex sample preparation which could potentially introduce analytical biases. Also, urinary ^1^H NMR spectroscopy has been applied in many other cohort studies and proposed as an objective method for dietary assessment^13^ potentially facilitating comparative studies in the future. We have used a pooled urine sample collection design which combined the last sample before bedtime with the first morning void sample of the following day, and we have shown in our preliminary work that this sample collection strategy has the advantage of reducing diurnal variations.^9, 70^

Our study has also some limitations. As in any observational study, there is the possibility of unmeasured residual confounding. In our analyses, we took into account a number of sociodemographic and lifestyle factors in childhood (e.g., socioeconomic status, ethnicity, sedentary behavior) that are associated with both diet quality and glycemic response. We did not have data available to control for energy intake. However, in all our models, we included BMI of the children, a measure strongly correlated to energy intake,^71^ and assessed ultra-processed food intake as proportion of total food intake. Moreover, the absence of heterogeneity across cohorts with different correlation structures and confounding patterns in their data^32^, provide evidence to support that unmeasured confounding is unlikely to have influenced the observed associations. Since the data collected are cross-sectional, and there is no temporality in the observed associations, further longitudinal studies examining metabolic and glycemic alterations in relation to diet quality are needed. Although ^1^H NMR spectroscopy had the advantage of improving the specificity of the quantitation and provided explicit metabolite identification, it limited the number of metabolites being measured and provided partial coverage of the urine metabolome. Absolute quantification of some metabolites with exchangable protons such as urea could also be negatively impacted by the solvent suppression methods required for ^1^H NMR spectroscopy of urine. Supplementing the current study with other complementary untargeted and targeted metabolomic approaches in future, such as mass spectrometry, would help enhance identification and robust quantification of urinary metabolites associated with diet quality in children.

In summary, this multi-center European study showed that urinary metabolic profiles related to food constituents (or their metabolic products), to amino acid and carbohydrate metabolism reflect adherence to the Mediterranean diet and ultra-processed food intake in childhood. Higher adherence to Mediterranean diet, lower ultra-processed food intake, and lower levels of the diet-related carbohydrate sucrose were associated with lower C-peptide levels, a marker of β-cell function. These results provide evidence to support efforts by public health authorities to recommend increased adherence to the Mediterranean diet and limiting UPF consumption in childhood. Further prospective studies examining the association of diet quality and related metabolomic profiles with C-peptide and other surrogates of insulin resistance are needed to replicate our findings.

## Data Availability

The HELIX data warehouse has been established as an accessible resource for collaborative research involving researchers external to the project. Access to HELIX data is based on approval by the HELIX Project Executive Committee and by the individual cohorts. Further details on the content of the data warehouse (data catalog) and procedures for external access are described on the project website (http://www.projecthelix.eu/index.php/es/data-inventory).

## Funding

The research leading to these results has received funding from the European Community’s Seventh Framework Programme (FP7/2007–2013) under grant agreement no 308333 – the HELIX project. The STOP project (http://www.stopchildobesity.eu/) received funding from the European Union’s Horizon 2020 research and innovation programme under Grant Agreement No. 774548. The STOP Consortium is coordinated by Imperial College London and includes 24 organisations across Europe, the United States and New Zealand. The content of this publication reflects only the views of the authors, and the European Commission is not liable for any use that may be made of the information it contains.

INMA data collections were supported by grants from the Instituto de Salud Carlos III, CIBERESP, and the Generalitat de Catalunya-CIRIT. KANC was funded by the grant of the Lithuanian Agency for Science Innovation and Technology (6-04-2014_31V-66). For a full list of funding that supported the EDEN cohort, refer to: Heude B et al. Cohort Profile: The EDEN mother-child cohort on the prenatal and early postnatal determinants of child health and development. Int J Epidemiol. 2016 Apr;45(2):353-63. The Norwegian Mother, Father and Child Cohort Study (MoBa) is supported by the Norwegian Ministry of Health and Care Services and the Ministry of Education and Research. The Rhea project was financially supported by European projects, and the Greek Ministry of Health (Program of Prevention of Obesity and Neurodevelopmental Disorders in Preschool Children, in Heraklion district, Crete, Greece: 2011–2014; ‘Rhea Plus’: Primary Prevention Program of Environmental Risk Factors for Reproductive Health, and Child Health: 2012–2015). Born in Bradford received funding from the Wellcome Trust (101597). Professor Wright and McEachan receive funding from the National Institute for Health Research Applied Research Collaboration for Yorkshire and Humber. The views expressed are those of the author(s) and not necessarily those of the NIHR or the Department of Health and Social Care. Dr. Maribel Casas received funding from Instituto de Salud Carlos III (Ministry of Economy and Competitiveness) (MS16/00128). Dr. Leda Chatzi was supported by NIH/NIHES R01 ES029944, R01ES030691, R01ES030364, R21 ES029681, R21 ES028903, and P30 ES007048-23. Dr. David Conti was supported by P01CA196569, R01CA140561, R01 ES016813, R01 ES029944, R01ES030691, R01ES030364. Dr. Nikos Stratakis was supported by NIH/NIHES R21 ES029681 and P30 ES007048-23, and NIH/NIDDK P30 DK048522-24. Dr. Hector Keun & Dr. Alexandros Siskos were also supported by the European Union’s Horizon 2020 research and innovation programme under grant agreement No 874583 (“ATHLETE”). Dr. Eleni Papadopoulou was supported by the Research Council of Norway, under the MILJØFORSK program (project number: 268465). Dr. Fernanda Rauber was supported by the Fundação de Amparo à Pesquisa do Estado de São Paulo 2016/14302-7 and 2018/19820-1. The CRG/UPF Proteomics Unit is part of the Spanish Infrastructure for Omics Technologies (ICTS OmicsTech) and it is a member of the ProteoRed PRB3 consortium which is supported by grant PT17/0019 of the PE I+D+i 2013-2016 from the Instituto de Salud Carlos III (ISCIII) and ERDF. ISGlobal acknowledges support from the Spanish Ministry of Science, Innovation and Universities, “Centro de Excelencia Severo Ochoa 2013-2017”, SEV-2012-0208, and “Secretaria d’Universitats i Recerca del Departament d’Economia i Coneixement de la Generalitat de Catalunya” (2017SGR595).

## Author Contributions

Nikos Stratakis (USC), Alexandros P. Siskos (Imperial), Eleni Papadopoulou (NIPH), Hector C. Keun (Imperial) and Leda Chatzi (USC) designed analytical and statistical methods. Alexandros P. Siskos, Chung-Ho E. Lau (Imperial), and Hector C. Keun generated and reviewed the primary metabolomics data. Nikos Stratakis, Anh N. Nguyen (Erasmus), and Katerina Margetaki (USC) analyzed the data. Nikos Stratakis, Alexandros P. Siskos, Eleni Papadopoulou, Hector C. Keun and Leda Chatzi wrote the first draft of the paper. Martine Vrijheid (ISGlobal) coordinated the HELIX project. Martine Vrijheid, Lea Maitre, and Oliver Robinson (ISGlobal) coordinated the HELIX data collection. All authors contributed to the data collection and/or interpretation of the results. All authors contributed to and approved the manuscript.

## Acknowledgments

The authors declare no conflict of interest. We acknowledge the input of the entire HELIX consortium. We are grateful to all the participating families in the five cohorts (BiB, EDEN, INMA, MoBa, KANC and RHEA cohorts), that took part in this study. We are equally grateful to all the fieldworkers for their dedication and efficiency in this study. A full roster of the INMA and RHEA study investigators can be found at http://www.proyectoinma.org/en/inma-project/inma-project-researchers/ and http://www.rhea.gr/en/about-rhea/the-rheateam/, respectively. The Born in Bradford study is only possible because of the enthusiasm and commitment of the participating children and parents. We are grateful to all the participants, health professionals and researchers who have made Born in Bradford happen. We are also grateful to all the participating families in Norway who take part in the on-going MoBa cohort study. We thank all the children and families participating in the EDEN-HELIX mother-child cohort. We are grateful to Joane Quentin, Lise Giorgis-Allemand, and Rémy Slama (EDEN study group) for their work on the HELIX project. We thank Sonia Brishoual, Angelique Serre and Michele Grosdenier (Poitiers Biobank, CRB BB-0033-00068, Poitiers, France) for biological sample management and Prof Frederic Millot (principal investigator), Elodie Migault, Manuela Boue and Sandy Bertin (Clinical Investigation Center, Inserm CIC1402, CHU de Poitiers, Poitiers, France) for planification and investigational actions. We are also grateful to Veronique Ferrand-Rigalleau, Celine Leger and Noella Gorry (CHU de Poitiers, Poitiers, France) for administrative assistance. We also acknowledge the commitment of the members of the EDEN Mother-Child Cohort Study Group: I. Annesi-Maesano, J.Y. Bernard, J. Botton, M.A. Charles, P. Dargent-Molina, B. de Lauzon-Guillain, P. Ducimetière, M. de Agostini, B. Foliguet, A. Forhan, X. Fritel, A. Germa, V. Goua, R. Hankard, M. Kaminski, B. Larroque, N. Lelong, J. Lepeule, G. Magnin, L. Marchand, C. Nabet, F Pierre, M.J. Saurel-Cubizolles, M. Schweitzer, O. Thiebaugeorges.

## Supplementary material

**Supplementary Table 1.**
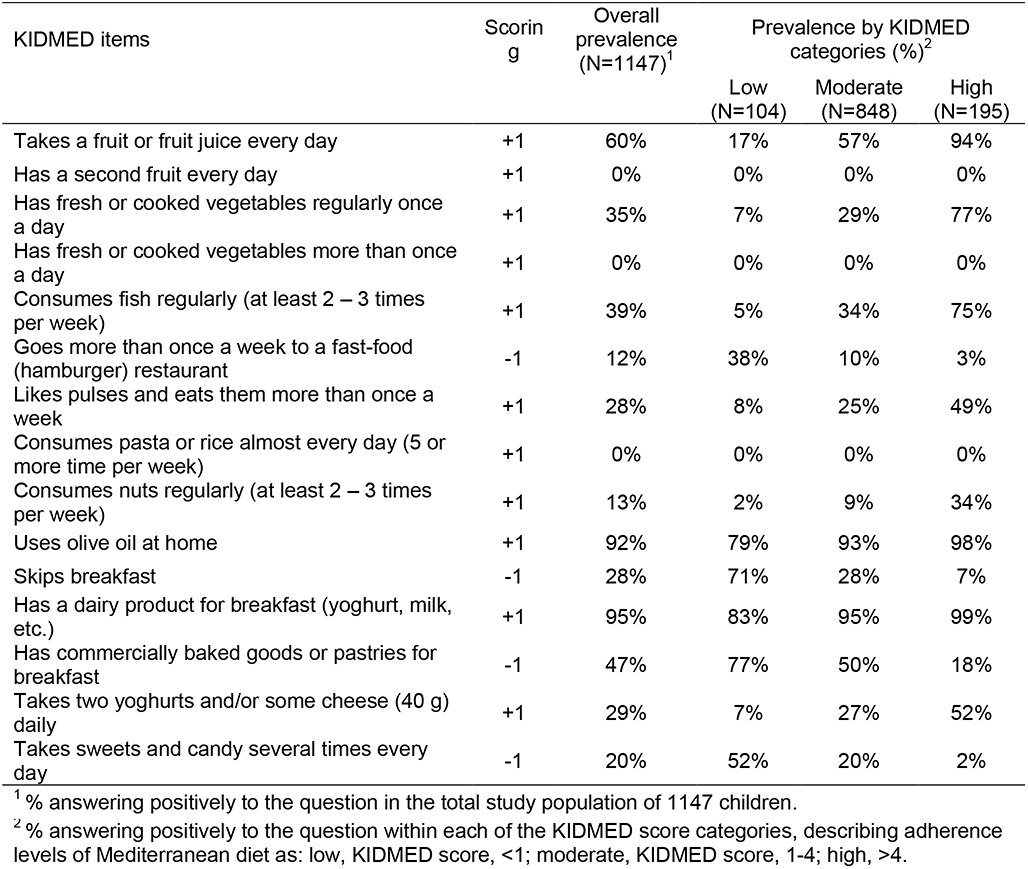
KIDMED items, and their scoring, used to assess adherence to the Mediterranean diet in HELIX children.

| KIDMED items | Scoring | Overall prevalence (N=1147)<sup>1</sup> | Prevalence by KIDMED categories (%)<sup>2</sup> |  |  |
| --- | --- | --- | --- | --- | --- |
|  |  |  | Low (N=104) | Moderate (N=848) | High (N=195) |
| Takes a fruit or fruit juice every day | +1 | 60% | 17% | 57% | 94% |
| Has a second fruit every day | +1 | 0% | 0% | 0% | 0% |
| Has fresh or cooked vegetables regularly once a day | +1 | 35% | 7% | 29% | 77% |
| Has fresh or cooked vegetables more than once a day | +1 | 0% | 0% | 0% | 0% |
| Consumes fish regularly (at least 2 – 3 times per week) | +1 | 39% | 5% | 34% | 75% |
| Goes more than once a week to a fast-food (hamburger) restaurant | -1 | 12% | 38% | 10% | 3% |
| Likes pulses and eats them more than once a week | +1 | 28% | 8% | 25% | 49% |
| Consumes pasta or rice almost every day (5 or more times per week) | +1 | 0% | 0% | 0% | 0% |
| Consumes nuts regularly (at least 2 – 3 times per week) | +1 | 13% | 2% | 9% | 34% |
| Uses olive oil at home | +1 | 92% | 79% | 93% | 98% |
| Skips breakfast | -1 | 28% | 71% | 28% | 7% |
| Has a dairy product for breakfast (yoghurt, milk, etc.) | +1 | 95% | 83% | 95% | 99% |
| Has commercially baked goods or pastries for breakfast | -1 | 47% | 77% | 50% | 18% |
| Takes two yoghurts and/or some cheese (40 g) daily | +1 | 29% | 7% | 27% | 52% |
| Takes sweets and candy several times every day | -1 | 20% | 52% | 20% | 2% |

<sup>1</sup> % answering positively to the question in the total study population of 1147 children.
<sup>2</sup> % answering positively to the question within each of the KIDMED score categories, describing adherence levels of Mediterranean diet as: low, KIDMED score, <1; moderate, KIDMED score, 1-4; high, >4.

**Supplementary Table 2.** Items included in the FFQ by ultra-processed food (UPF) inclusion and information regarding the extent and purpose of food processing.

| Food item in the FFQ<sup>1</sup> | Description | Information on extent and purpose of food processing<sup>2</sup> |
| --- | --- | --- |
| **Foods in the UPF group** |  |  |
| Cookies | Plain biscuits and cookies | Assumption that most of these products are industrially made formulations of ingredients that result from a series of industrial processes, many requiring sophisticated equipment and technology. Such processes include the fractioning of whole foods into substances, chemical modifications of these substances, assembly of unmodified and modified food substances using industrial techniques such as extrusion, moulding and pre-frying, frequent application of additives whose function is to make the final product palatable or hyper-palatable ('cosmetic additives'), and sophisticated packaging, usually with synthetic materials.<br>The used ingredients often include sugar, oils and fats, and salt, generally in combination; substances that are sources of energy and nutrients but of no or rare culinary use such as high fructose corn syrup, hydrogenated or interesterified oils, and protein isolates; cosmetic additives such as flavours, flavour enhancers, colours, emulsifiers, sweeteners, thickeners, and anti-foaming, bulking, carbonating. |
| Pastries | Buns, cakes, pastries, croissants, donuts, halva |  |
| Sugar-Sweetened Beverages | Sugar-sweetened soft/fizzy drinks (eg coca cola, fanta, ribena) |  |
| Artificially-sweetened and low-sugar beverages | Artificially sweetened/low sugar/sugar free soft drinks (eg. Diet fanta) |  |
| Cold meat cuts | Sausages, bacon, chorizo, salami (in slices) |  |
| Ham | Ham (in slices) |  |
| Dairy deserts | Dairy desert/ milky pudding, ice cream |  |
| Sugar sweetened breakfast cereals | Sugar-sweetened cereals for children |  |
| Other breakfast cereals | Other breakfast cereals/porridge |  |
| Crispbreads and rusks | Rusks, crispy bread, rice and corn cakes |  |
| Chocolate | Chocolate (bars, bonbon, spreads, cacao) |  |
| Sweets | Sweets, gummy bears |  |
| Margarine | Margarine |  |
| Dressings | Mayonnaise, ketchup, dressings |  |
| Salty snacks | Crisps, salty snacks, pipas |  |
| **Food not in the UPF group** |  |  |
| Red meat | Red meat (beef, pork lamb, steak, burger, chop, meatballs, small curry, zeppelin meat dumpling, minced meat) | Assumption that most of these products are whole or in the form of steaks, fillets and other cuts, fresh or chilled or frozen. |
| Poultry | Poultry (chicken, turkey, burger, nuggets, curry) | Assumption that most of these products are whole or in the form of steaks, fillets and other cuts, fresh or chilled or frozen. |
| White fish | Lean/white fish (flat fish, perch, fish cakes) | Assumption that most of these products are whole or in the form of steaks, fillets and other cuts, fresh or chilled or frozen. |
| Oily fish | Fatty/oily fish (sardines, salmon, mackerel, tuna, trout, carp) | Assumption that most of these products are whole or in the form of steaks, fillets and other cuts, fresh or chilled or frozen. |
| Seafood | Seafood (shellfish, squids, shrimps) | Assumption that most of these products are whole or in the form of steaks, fillets and other cuts, fresh or chilled or frozen. |
| Canned fish | Canned fish (tuna, sardines, sprat) | Assumption that most of these products are processed in order to increase the durability of fresh foods and may contain additives that prolong product duration, protect original properties or prevent proliferation of microorganisms. |
| Egg | Egg (fried, scrambled, boiled, omelet) | Assumption that most of these products are made of fresh or chilled eggs. |
| Cheese | Cheese (eg cheddar, brie, feta, cures cheese, brown cheese, curd, cream cheese) | Assumption that most of these products are processed in order to increase the durability and make them more enjoyable by modifying or enhancing their sensory qualities. These products may contain additives that prolong product duration, protect original properties or |
|  |  | prevent proliferation of microorganisms. |
| Milk | Milk (skimmed, semiskimmed, full fat milk) | Assumption that most of these products are not artificially flavored |
| Probiotic yoghurt | Pro-biotic yoghurt (eg. Actimel, kefir) | Assumption that most of these products are not artificially flavored |
| Yoghurt | Yoghurt (excluding pro-biotic) | Assumption that most of these products are not artificially flavored |
| Fruits | Fruits | Assumption that most of these products are fresh, squeezed, chilled, frozen fruits. |
| Canned fruit | Canned fruit with syrup/nectars | Assumption that most of these products are processed in order to increase the durability of fresh foods and may contain additives that prolong product duration, protect original properties or prevent proliferation of microorganisms. |
| Juice | Natural/pure/freshly squeezed fruit juice | Assumption that most of these products do not contain flavorings and sweeteners |
| Dry fruits | Dried fruit (raisins, currant, dry apricots, prunes, figs) | Assumption that most of these products are dried fruits. |
| Cooked vegetables | Cooked or pureed vegetables (no potatoes) | Assumption that most of these products are of fresh, squeezed, chilled, frozen, or dried leafy and root vegetables or from canned or bottled vegetables. |
| Potatoes | Potatoes (boiled, baked, mashed) not chips/fries | Assumption that most of these products are homemade from fresh potatoes. |
| Fries | French fries (1 portion) | Assumption that most of these products are homemade from fresh potatoes. |
| Vegetables | Raw vegetables | Assumption that most of these products are fresh, squeezed, chilled, frozen, or dried leafy and root vegetables. |
| Nuts | Nuts (almonds, walnuts) | Assumption that most of these products are nuts with or without salt or sugar. |
| Pulses | Pulses (chickpeas, lentils, peas, beans) | Assumption that most of these products are fresh or canned/bottled legumes. |
| Dark bread | Whole grain /dark bread/ '50/50" | Assumption that most of these products are fresh bread that contain different types of flour, water and yeast. |
| White bread | White bread | Assumption that most of these products are fresh bread that contain different types of flour, water and yeast. |
| Rice & pasta | Rice and pasta | Assumption that most of these products are grains and past made of flour. |
| Sugar and other sweeteners | Sugar, honey, jam | Assumption that most of these products are sugar from cane or beet, honey extracted from combs and jam made from fruit, sugar, water and other additives that prolong product duration. |
| Butter | Butter or ghee | Assumption that most of these products are from milk. |
| Olive oil | Olive oil for cooking or seasoning | Assumption that most of these products are vegetable oils crushed from olives. |
| Other oils | Other vegetable oils (canola oil) | Assumption that most of these products are vegetable oils crushed from seeds and nuts. |

<sup>1</sup> Every item corresponds to one question in the FFQ.
<sup>2</sup> NOVA food groups: definition according to the extent and purpose of food processing from Monteiro CA et al. Ultra-processed foods: what they are and how to identify them. Public Health Nutrition (2019), Volume 22, Issue 5, pp. 936-94.

**Supplementary Table 3.** Intakes of food groups (in servings/week) by categories of the KIDMED score^1^.

|  | KIDMED categories<sup>2</sup> |  |  | P value<sup>3</sup> |
| --- | --- | --- | --- | --- |
|  | Low (N=104) | Moderate (N=848) | High (N=195) |  |
| Vegetables | 4.5 (4.3) | 7.6 (5.7) | 13.5 (7.1) | <0.001 |
| Fruits | 6.3 (5.7) | 11.2 (7.9) | 16.8 (7.8) | <0.001 |
| Fish and seafood | 1.3 (0.9) | 2.5 (2.2) | 4 (2.7) | <0.001 |
| Meat | 8.1 (4.2) | 7.9 (4.5) | 8 (4.1) | 0.67 |
| Dairy products | 14.8 (8.7) | 21.1 (11.2) | 24.4 (12.1) | <0.001 |
| Pulses | 0.7 (0.9) | 1.2 (1.2) | 1.8 (1.3) | <0.001 |
| Bread & Cereals | 9.6 (8.2) | 11 (7.3) | 13.1 (7.1) | <0.001 |
| Eggs | 1.8 (1.5) | 1.8 (1.6) | 2 (1.8) | 0.33 |
| Nuts | 0.4 (0.5) | 0.7 (1.3) | 1.6 (1.9) | <0.001 |
| Potatoes | 3.8 (2.7) | 3.7 (2.7) | 3.7 (2.7) | 0.46 |
| Bakery products | 7.1 (5.5) | 5.3 (5) | 3.6 (3) | <0.001 |
| Added fats & oils | 2.4 (3.2) | 1.8 (2.9) | 1.5 (2.5) | 0.002 |
| Salty snacks | 1.7 (1.9) | 1.4 (2) | 1.4 (1.7) | 0.09 |
| Sweets | 11.6 (8.8) | 7.5 (6) | 6.7 (4.5) | <0.001 |
| Beverages | 2.8 (5.5) | 1.2 (3.1) | 1 (2.2) | 0.003 |
| Dressings | 1.7 (1.7) | 2.3 (2.8) | 1.9 (1.9) | 0.24 |

<sup>1</sup> Values are mean (SD).
<sup>2</sup> KIDMED score categories were defined as follows: low, <1; moderate, 1-4; high, >4.
<sup>3</sup> P-value for differences across KIDMED score categories were derived using Kruskal-Wallis test.

**Supplementary Table 4.**
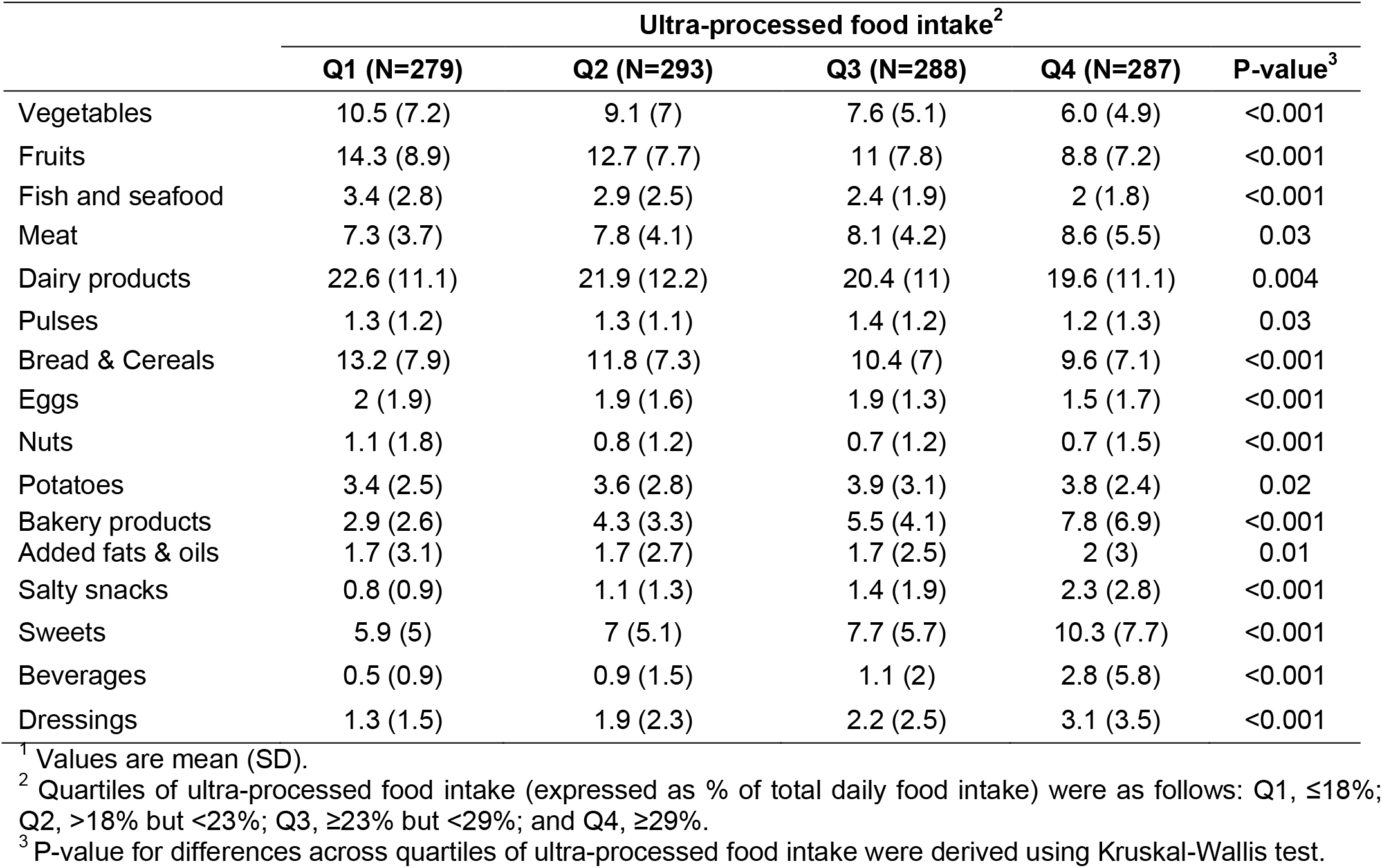
Intakes of food groups (in servings/week) by quartiles of ultra-processed food intake^1^.

|  | Ultra-processed food intake<sup>2</sup> |  |  |  | P-value<sup>3</sup> |
| --- | --- | --- | --- | --- | --- |
|  | Q1 (N=279) | Q2 (N=293) | Q3 (N=288) | Q4 (N=287) |  |
| Vegetables | 10.5 (7.2) | 9.1 (7) | 7.6 (5.1) | 6.0 (4.9) | <0.001 |
| Fruits | 14.3 (8.9) | 12.7 (7.7) | 11 (7.8) | 8.8 (7.2) | <0.001 |
| Fish and seafood | 3.4 (2.8) | 2.9 (2.5) | 2.4 (1.9) | 2 (1.8) | <0.001 |
| Meat | 7.3 (3.7) | 7.8 (4.1) | 8.1 (4.2) | 8.6 (5.5) | 0.03 |
| Dairy products | 22.6 (11.1) | 21.9 (12.2) | 20.4 (11) | 19.6 (11.1) | 0.004 |
| Pulses | 1.3 (1.2) | 1.3 (1.1) | 1.4 (1.2) | 1.2 (1.3) | 0.03 |
| Bread & Cereals | 13.2 (7.9) | 11.8 (7.3) | 10.4 (7) | 9.6 (7.1) | <0.001 |
| Eggs | 2 (1.9) | 1.9 (1.6) | 1.9 (1.3) | 1.5 (1.7) | <0.001 |
| Nuts | 1.1 (1.8) | 0.8 (1.2) | 0.7 (1.2) | 0.7 (1.5) | <0.001 |
| Potatoes | 3.4 (2.5) | 3.6 (2.8) | 3.9 (3.1) | 3.8 (2.4) | 0.02 |
| Bakery products | 2.9 (2.6) | 4.3 (3.3) | 5.5 (4.1) | 7.8 (6.9) | <0.001 |
| Added fats & oils | 1.7 (3.1) | 1.7 (2.7) | 1.7 (2.5) | 2 (3) | 0.01 |
| Salty snacks | 0.8 (0.9) | 1.1 (1.3) | 1.4 (1.9) | 2.3 (2.8) | <0.001 |
| Sweets | 5.9 (5) | 7 (5.1) | 7.7 (5.7) | 10.3 (7.7) | <0.001 |
| Beverages | 0.5 (0.9) | 0.9 (1.5) | 1.1 (2) | 2.8 (5.8) | <0.001 |
| Dressings | 1.3 (1.5) | 1.9 (2.3) | 2.2 (2.5) | 3.1 (3.5) | <0.001 |

<sup>1</sup> Values are mean (SD).<sup>2</sup> Quartiles of ultra-processed food intake (expressed as % of total daily food intake) were as follows: Q1, ≤18%; Q2, >18% but <23%; Q3, ≥23% but <29%; and Q4, ≥29%.<sup>3</sup> P-value for differences across quartiles of ultra-processed food intake were derived using Kruskal-Wallis test.

**Supplementary Table 5.** Associations of KIDMED score with urinary metabolites in childhood^1^.

| Urinary metabolite | Compound Class | Percent change (95% CI) | P-value | Q-value |
| --- | --- | --- | --- | --- |
| scyllo-inositol | Alcohols and polyols | 0.23 (-4.98, 5.64) | 0.95 | 0.95 |
| N-methylnicotinic acid | Alkaloids and derivatives | 4.71 (1.67, 7.65) | <0.001 | 0.03 |
| N-methylpicolinic acid | Alkaloids and derivatives | 3.99 (-2.44, 10.71) | 0.23 | 0.48 |
| 2-hydroxyisobutyrate | Alpha hydroxy acids and derivatives | 1.16 (-0.15, 2.28) | 0.09 | 0.29 |
| Lactate<sup>2</sup> | Alpha hydroxy acids and derivatives | -0.92 (-2.96, 1.15) | 0.38 | 0.63 |
| Dimethylamine | Amines | 1.86 (-0.91, 4.55) | 0.2 | 0.44 |
| Trimethylamine | Amines | 2.09 (0.33, 3.67) | 0.02 | 0.11 |
| 3-aminoisobutyrate | Amino acids, peptides, and analogues | 1.39 (-2.1, 4.85) | 0.45 | 0.67 |
| 5-oxoproline | Amino acids, peptides, and analogues | 0.23 (-2.12, 2.72) | 0.83 | 0.89 |
| Alanine | Amino acids, peptides, and analogues | -1.37 (-2.86, 0.23) | 0.1 | 0.29 |
| Creatine | Amino acids, peptides, and analogues | 0.93 (-3.03, 5.08) | 0.65 | 0.84 |
| Glutamine | Amino acids, peptides, and analogues | -0.92 (-4.15, 2.27) | 0.55 | 0.73 |
| Glycine | Amino acids, peptides, and analogues | -1.14 (-2.93, 0.71) | 0.23 | 0.48 |
| Isoleucine | Amino acids, peptides, and analogues | 2.09 (-0.86, 5.18) | 0.17 | 0.41 |
| Leucine | Amino acids, peptides, and analogues | 0.23 (-0.93, 1.47) | 0.67 | 0.84 |
| Lysine | Amino acids, peptides, and analogues | -0.23 (-2.59, 2.06) | 0.81 | 0.89 |
| Proline betaine | Amino acids, peptides, and analogues | 6.66 (-2.04, 16.05) | 0.14 | 0.37 |
| Tyrosine | Amino acids, peptides, and analogues | 2.33 (0.29, 4.2) | 0.02 | 0.13 |
| Valine | Amino acids, peptides, and analogues | 0.69 (-0.48, 1.72) | 0.27 | 0.51 |
| 3-hydroxybutyrate/3-aminoisobutyrate<sup>3</sup> | Amino acids, peptides, and analogues | 0.93 (-4.14, 6.16) | 0.74 | 0.88 |
| Trimethylamine N-oxide | Aminoxides | 5.44 (-0.31, 11.66) | 0.06 | 0.23 |
| 3-Indoxylsulfate | Arylsulfates | 3.51 (-0.07, 7.07) | 0.06 | 0.21 |
| p-cresol sulfate | Arylsulfates | 3.28 (0.04, 6.83) | 0.05 | 0.21 |
| Hippurate | Benzoic acids and derivatives | 5.93 (3.39, 8.55) | <0.001 | <0.001 |
| 3-hydroxyisobutyrate | Beta hydroxy acids and derivatives | -0.46 (-2.24, 1.16) | 0.53 | 0.73 |
| Glucose | Carbohydrates and carbohydrate conjugates | -0.23 (-1.94, 1.58) | 0.83 | 0.89 |
| N-acetyl neuraminic acid | Carbohydrates and carbohydrate conjugates | 0.46 (-2.68, 3.64) | 0.79 | 0.89 |
| Sucrose | Carbohydrates and carbohydrate conjugates | -5.81 (-9.87, -1.67) | 0.01 | 0.05 |
| Acetate | Carboxylic acids | 3.51 (0.95, 6.07) | 0.01 | 0.05 |
| Formate | Carboxylic acids | 1.86 (-0.38, 3.94) | 0.11 | 0.31 |
| Carnitine<sup>4</sup> | Carnitines | -2.28 (-6.45, 1.97) | 0.28 | 0.51 |

**Supplementary Table 5- Continued**
| Urinary metabolite | Compound Class | Percent change (95% CI) | P value | Q value |
| --- | --- | --- | --- | --- |
| Succinate | Dicarboxylic acids and derivatives | 3.51 (-3.61, 11.28) | 0.34 | 0.58 |
| 3-hydroxyisovalerate | Hydroxy fatty acids | 0.23 (-1.2, 1.85) | 0.69 | 0.84 |
| Acetone | Ketones | 2.33 (-0.98, 5.6) | 0.17 | 0.41 |
| *N*1-methyl-nicotinamide | Nicotinamide | 1.16 (-1.68, 4.23) | 0.41 | 0.63 |
| *N*-methyl-2-pyridone-5-carboxamide | Nicotinamides | 0.46 (-4.73, 5.98) | 0.86 | 0.9 |
| Urea | Organic carbonic acids and derivatives | 2.33 (1.04, 3.47) | <0.001 | 0.01 |
| Taurine | Organosulfonic acids | 1.62 (-2.84, 6.51) | 0.46 | 0.67 |
| *p*-hydroxyphenylacetate | Phenols | 1.62 (0, 3.07) | 0.05 | 0.21 |
| Pantothenic acid | Polyols | 1.62 (0.55, 2.55) | <0.001 | 0.03 |
| 4-deoxyerythronic acid | Sugar acids and derivatives | 0.93 (-0.68, 2.53) | 0.26 | 0.51 |
| 4-deoxythreonic acid | Sugar acids and derivatives | 0.69 (-1.04, 2.59) | 0.41 | 0.63 |
| Citrate | Tricarboxylic acids and derivatives | 0.00 (-2.23, 2.47) | 0.94 | 0.95 |

Abbreviations: KIDMED score, Mediterranean Diet Quality Index for children and adolescents.
<sup>1</sup> Effect estimates are percent changes (95% CI) in metabolite levels (in log<sub>10</sub> μmol/mmol of creatinine) per 1-unit increase in the KIDMED score and were derived from linear regression models adjusted for maternal age, maternal education level, maternal pre-pregnancy BMI, family affluence status, child sex, child age, child BMI, child sedentary behavior, child ethnicity, and a cohort indicator.
<sup>2</sup> Threonine could also contribute as a minor component in the signal for lactate
<sup>3</sup> Overlapping signal between 3-hydroxybutyrate/3-aminoisobutyrate
<sup>4</sup> Choline could also contribute as a minor component in the signal of carnitine

**Supplementary Table 6.** Associations of ultra-processed food consumption with urinary metabolites in childhood^1^.

| Urinary metabolite | Compound Class | Percent change (95% CI) | P-value | Q-value |
| --- | --- | --- | --- | --- |
| scyllo-inositol | Alcohols and polyols | 0.69 (-4.46, 5.98) | 0.814 | 0.853 |
| N-methylnicotinic acid | Alkaloids and derivatives | -3.84 (-6.59, -1.22) | 0.005 | 0.035 |
| N-methylpicolinic acid | Alkaloids and derivatives | -1.83 (-7.71, 4.45) | 0.561 | 0.71 |
| 2-hydroxyisobutyrate | Alpha hydroxy acids and derivatives | -1.6 (-2.65, -0.33) | 0.012 | 0.074 |
| Lactate<sup>2</sup> | Alpha hydroxy acids and derivatives | -0.92 (-3, 1.03) | 0.332 | 0.528 |
| Dimethylamine | Amines | 0.69 (-1.99, 3.29) | 0.648 | 0.774 |
| Trimethylamine | Amines | -0.46 (-1.97, 1.23) | 0.642 | 0.774 |
| 3-aminoisobutyrate | Amino acids, peptides, and analogues | -3.17 (-6.32, 0.17) | 0.063 | 0.225 |
| 3-hydroxybutyrate/3-aminoisobutyrate<sup>3</sup> | Amino acids, peptides, and analogues | -2.95 (-7.69, 1.99) | 0.235 | 0.422 |
| 5-oxoproline | Amino acids, peptides, and analogues | -1.83 (-4.15, 0.47) | 0.117 | 0.255 |
| Alanine | Amino acids, peptides, and analogues | -0.46 (-2.06, 0.99) | 0.484 | 0.676 |
| Creatine | Amino acids, peptides, and analogues | -0.92 (-4.68, 3.12) | 0.668 | 0.776 |
| Glutamine | Amino acids, peptides, and analogues | -0.69 (-3.71, 2.6) | 0.708 | 0.781 |
| Glycine | Amino acids, peptides, and analogues | 0 (-1.8, 1.81) | 0.991 | 0.991 |
| Isoleucine | Amino acids, peptides, and analogues | -2.5 (-5.21, 0.44) | 0.096 | 0.255 |
| Leucine | Amino acids, peptides, and analogues | -1.37 (-2.51, -0.2) | 0.022 | 0.093 |
| Lysine | Amino acids, peptides, and analogues | 0 (-2.34, 2.21) | 0.936 | 0.958 |
| Proline betaine | Amino acids, peptides, and analogues | -2.95 (-10.61, 5.53) | 0.491 | 0.676 |
| Tyrosine | Amino acids, peptides, and analogues | -2.95 (-4.67, -1.03) | 0.002 | 0.025 |
| Valine | Amino acids, peptides, and analogues | -1.6 (-2.62, -0.51) | 0.004 | 0.032 |
| Trimethylamine N-oxide | Aminoxides | -1.83 (-7.18, 3.73) | 0.503 | 0.676 |
| 3-Indoxylsulfate | Arylsulfates | -2.28 (-5.53, 1.07) | 0.179 | 0.35 |
| p-cresol sulfate | Arylsulfates | -2.95 (-6, 0.23) | 0.069 | 0.228 |
| Hippurate | Benzoic acids and derivatives | -5.59 (-7.78, -3.28) | <0.001 | <0.001 |
| 3-hydroxyisobutyrate | Beta hydroxy acids and derivatives | -1.37 (-2.96, 0.34) | 0.119 | 0.255 |
| Glucose | Carbohydrates and carbohydrate conjugates | -1.14 (-2.84, 0.57) | 0.188 | 0.352 |
| N-acetyl neuraminic acid | Carbohydrates and carbohydrate conjugates | 0.69 (-2.43, 3.77) | 0.694 | 0.781 |
| Sucrose | Carbohydrates and carbohydrate conjugates | 8.39 (3.9, 13.1) | <0.001 | 0.003 |
| Acetate | Carboxylic acids | -1.83 (-4.1, 0.66) | 0.154 | 0.316 |
| Formate | Carboxylic acids | -1.83 (-3.83, 0.26) | 0.086 | 0.255 |
| Carnitine<sup>4</sup> | Carnitines | -2.28 (-6.3, 1.95) | 0.288 | 0.476 |

**Supplementary Table 6- Continued**
| Urinary metabolite | Compound Class | Percent change (95% CI) | P-value | Q-value |
| --- | --- | --- | --- | --- |
| Succinate | Dicarboxylic acids and derivatives | -2.73 (-9.33, 4.35) | 0.44 | 0.652 |
| 3-hydroxyisovalerate | Hydroxy fatty acids | -1.6 (-3.08, -0.17) | 0.029 | 0.112 |
| Acetone | Ketones | -2.5 (-5.53, 0.6) | 0.113 | 0.255 |
| *N*1-methyl-nicotinamide | Nicotinamide | -1.14 (-3.95, 1.71) | 0.423 | 0.65 |
| *N*-methyl-2-pyridone-5-carboxamide | Nicotinamides | 0.69 (-4.35, 6.16) | 0.773 | 0.831 |
| Urea | Organic carbonic acids and derivatives | -2.73 (-3.76, -1.5) | <0.001 | <0.001 |
| Taurine | Organosulfonic acids | -5.38 (-9.52, -1.03) | 0.016 | 0.076 |
| *p*-hydroxyphenylacetate | Phenols | -0.46 (-1.91, 1.04) | 0.552 | 0.71 |
| Pantothenic acid | Polyols | -0.92 (-1.78, 0.15) | 0.095 | 0.255 |
| 4-deoxyerythronic acid | Sugar acids and derivatives | -0.92 (-2.38, 0.7) | 0.282 | 0.476 |
| 4-deoxythreonic acid | Sugar acids and derivatives | -2.05 (-3.84, -0.41) | 0.016 | 0.076 |
| Citrate | Tricarboxylic acids and derivatives | -1.83 (-4.05, 0.46) | 0.117 | 0.255 |

Abbreviations: UPF, ultra-processed food intake.
<sup>1</sup> Effect estimates are percent changes (95% CI) in metabolite levels (in log<sub>10</sub> μmol/mmol of creatinine) per 5% increase in daily UPF intake and were derived from linear regression models adjusted for maternal age, maternal education level, maternal pre-pregnancy BMI, family affluence status, child sex, child age, child BMI, child sedentary behavior, child ethnicity, and a cohort indicator.
<sup>2</sup> Threonine could also contribute as a minor component in the signal for lactate
<sup>3</sup> Overlapping signal between 3-hydroxybutyrate/3-aminoisobutyrate
<sup>4</sup> Choline could also contribute as a minor component in the signal of carnitine

**Supplementary Table 7.**
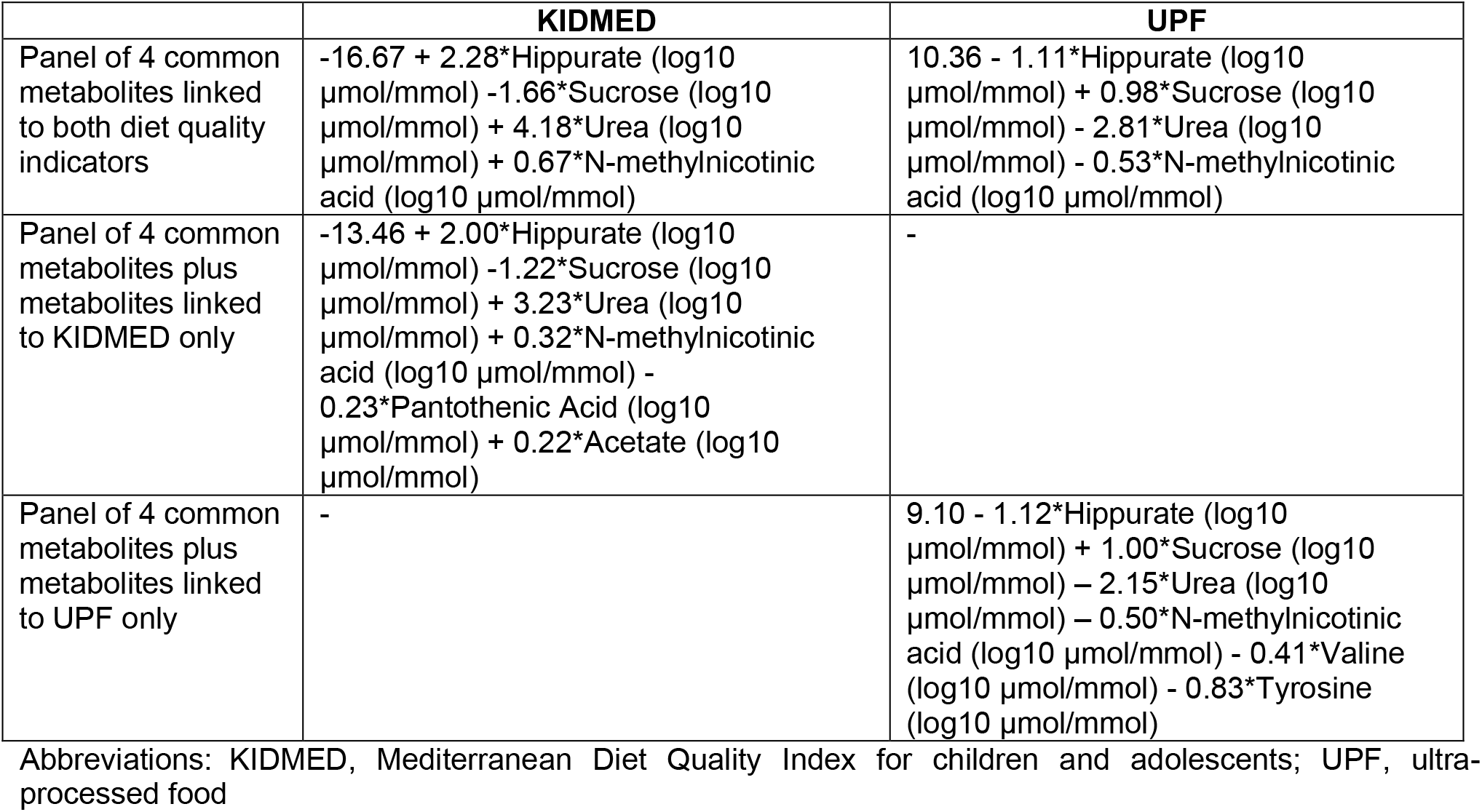
Regression formulas (scores) for predicting diet quality in childhood based on panels of urinary metabolites (creatinine-normalized concentrations, μmol/mmol of creatinine)

**Supplementary Table 8.** Interaction of diet quality indicators in association to Cpeptide concentration in childhood^1^.

|  | **C-peptide** |
| --- | --- |
|  | **Percent change (95% CI)** |
| KIDMED score (UPF intake, % of total daily food intake)<sup>2</sup> |  |
| KIDMED ≤4, UPF <23% | *Ref.* |
| KIDMED ≤4, UPF ≥23% | 20.9 (-3.9, 52.0) |
| KIDMED >4, UPF <23% | -16.2 (-39.8, 16.5) |
| KIDMED >4, UPF ≥23% | 21.3 (-29.6, 109.1) |
| P value for interaction | 0.57 |

Abbreviations: KIDMED, Mediterranean Diet Quality Index for children and adolescents; UPF, ultra-processed food
<sup>1</sup> Effect estimates represent percent changes in log-2 transformed C-peptide levels and their 95% CIs per combination of the KIDMED score and median-based UPF intake and were derived from linear regression models adjusted for maternal age, maternal education level, maternal pre-pregnancy BMI, family affluence status, child sex, child age, child BMI, child sedentary behavior, child ethnicity, postprandial interval, and a cohort indicator.
<sup>2</sup> The following number of children were included for each combination of the KIDMED score and UPF intake: 422 children with low/moderate KIDMED score (≤4) and UPF intake below the median population intake (<23%); 530 children with low/moderate KIDMED score and UPF intake above the median population intake (≥23%); 150 children with high KIDMED score and UPF intake below the median population intake (≥23%); and 45 children with high KIDMED score and UPF intake above the median population intake.

**Supplementary Table 9.** Associations of diet quality with C-peptide concentration in childhood after stratifying by sex and by weight status, respectively^1^.

|  | KIDMED score | UPF intake |
| --- | --- | --- |
| **Sex** |  |  |
| Boys (N=626) | -7.5 (-14.8, 0.5) | 7.5 (-0.8, 16.5) |
| Girls (N=521) | -8.8 (-16.4, -0.5) | 11.5 (2.1, 21.8) |
| P-interaction | 0.80 | 0.53 |
| **Weight status<sup>2</sup>** |  |  |
| Normal weight (N=906) | -9.5 (-15.5, -3.0) | 10.5 (3.2, 18.4) |
| Overweight/Obese (N=237) | -4.5 (-16.7, 9.5) | 4.6 (-7.1, 18.9) |
| P-interaction | 0.48 | 0.45 |

Abbreviations: KIDMED, Mediterranean Diet Quality Index for children and adolescents; UPF, ultra-processed food
<sup>1</sup> Effect estimates represent percent changes in log-2 transformed C-peptide levels and their 95% CIs per 1-unit increase in the KIDMED score or per 5% increase in daily UPF intake and were derived from linear regression models adjusted for maternal age, maternal education level, maternal pre-pregnancy BMI, family affluence status, child sex (for models not stratified by sex), child age, child BMI (for models not stratified by weight status), child sedentary behavior, child ethnicity, postprandial interval, and a cohort indicator.
<sup>2</sup> Categories of normal weight and overweight/obese were derived using the International Obesity Taskforce criteria.

**Supplementary Table 10.** Associations between urinary metabolites (μmol/mmol of creatinine) linked to diet quality and C-peptide in childhood^1^.

|  | C-peptide |
| --- | --- |
| **Metabolites scores based on panels of metabolites, per SD** |  |
| Score for KIDMED based on the 4 metabolites linked to both KIDMED and UPF<sup>2</sup> | -11.72 (-20.19, -2.34) |
| Score for KIDMED based on the 4 metabolites linked to both KIDMED and UPF plus metabolites linked to KIDMED only<sup>3</sup> | -11.91 (-20.36, -2.55) |
| Score for UPF based on the 4 metabolites linked to both KIDMED and UPF<sup>4</sup> | 11.90 (1.15, 23.79) |
| Score for UPF based on the 4 metabolites linked to both KIDMED and UPF plus metabolites linked to UPF only<sup>5</sup> | 11.66 (0.93, 23.52) |
| **Individual urinary metabolites, per log10 μmol/mmol** | **Percent change (95% CI)** |
| Acetate | 21 (-14, 70.1) |
| Hippurate | -13.2 (-38.4, 22.3) |
| N-methylnicotinic acid | 7.8 (-19.7, 44.7) |
| Pantothenic acid | 25.8 (-46.3, 194.8) |
| Sucrose | 24.7 (2.8, 51.3) |
| Tyrosine | -7.6 (-40.4, 43.5) |
| Urea | 17.5 (-42, 138.4) |
| Valine | 31.9 (-38.9, 185.1) |

Abbreviations: KIDMED, Mediterranean Diet Quality Index for children and adolescents; UPF, ultra-processed food
<sup>1</sup> Effect estimates derived from linear regression models represent percent changes in log-2 transformed C-peptide levels and their 95% CIs per log10 μmol/mmol increase in individual metabolite concentrations and SD increase in metabolite scores.
<sup>2</sup> Score= -16.67 + 2.28\*Hippurate (log10 μmol/mmol) -1.66\*Sucrose (log10 μmol/mmol) + 4.18\*Urea (log10 μmol/mmol) + 0.67\*N-methylnicotinic acid (log10 μmol/mmol)
<sup>3</sup> Score= -13.46 + 2.00\*Hippurate (log10 μmol/mmol) -1.22\*Sucrose (log10 μmol/mmol) + 3.23\*Urea (log10 μmol/mmol) + 0.32\*N-methylnicotinic acid (log10 μmol/mmol) - 0.23\*Pantothenic Acid (log10 μmol/mmol) + 0.22\*Acetate (log10 μmol/mmol)
<sup>4</sup> Score= 10.36 - 1.11\*Hippurate (log10 μmol/mmol) + 0.98\*Sucrose (log10 μmol/mmol) - 2.81\*Urea (log10 μmol/mmol) - 0.53\*N-methylnicotinic acid (log10 μmol/mmol)
<sup>5</sup> Score= 9.10 - 1.12\*Hippurate (log10 μmol/mmol) + 1.00\*Sucrose (log10 μmol/mmol) - 2.15\*Urea (log10 μmol/mmol) - 0.50\*N-methylnicotinic acid (log10 μmol/mmol) - 0.41\*Valine (log10 μmol/mmol) - 0.83\*Tyrosine (log10 μmol/mmol)

**Supplementary Figure 1.**
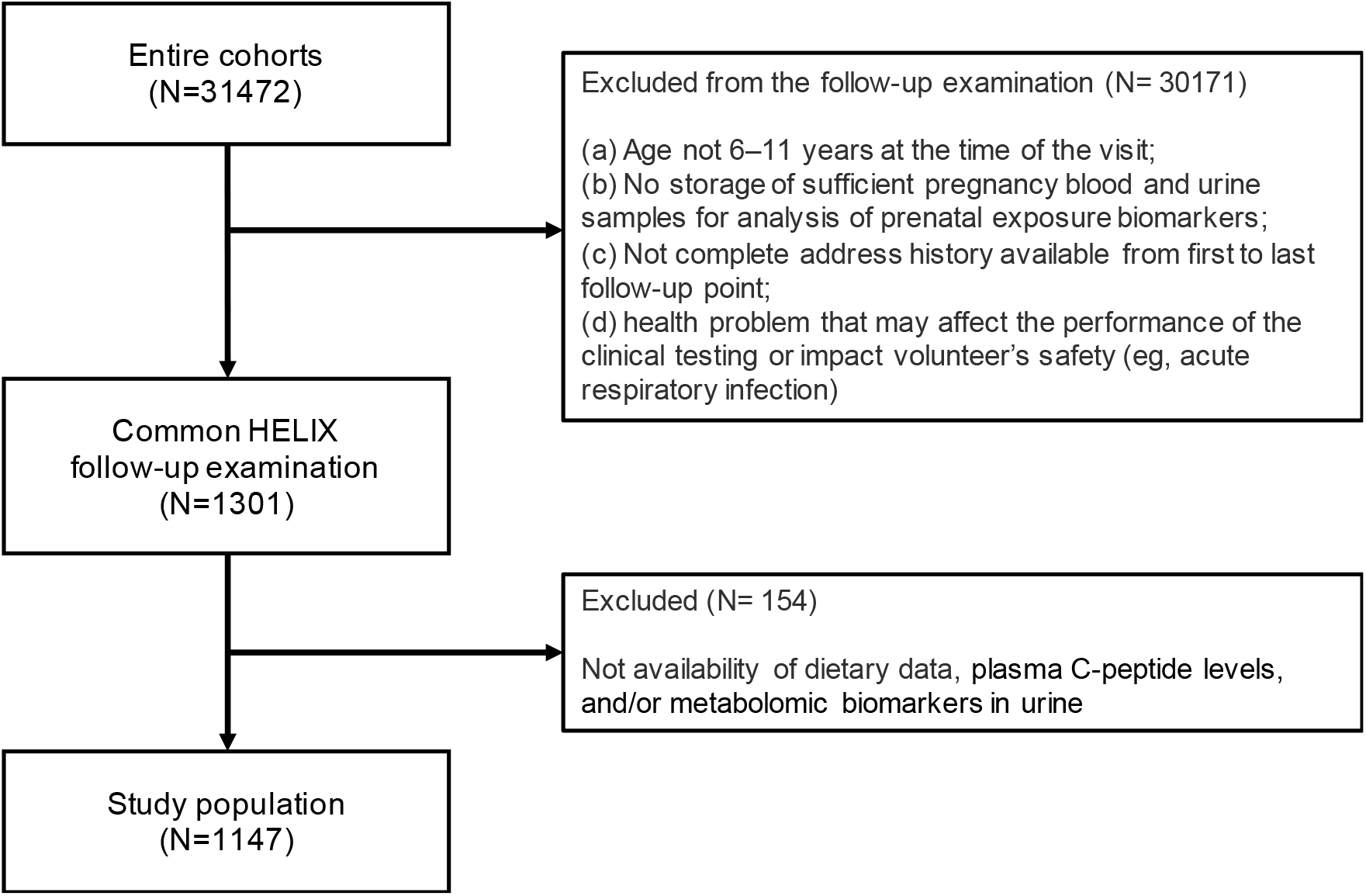
Participant flowchart.

**Supplementary Figure 2.**
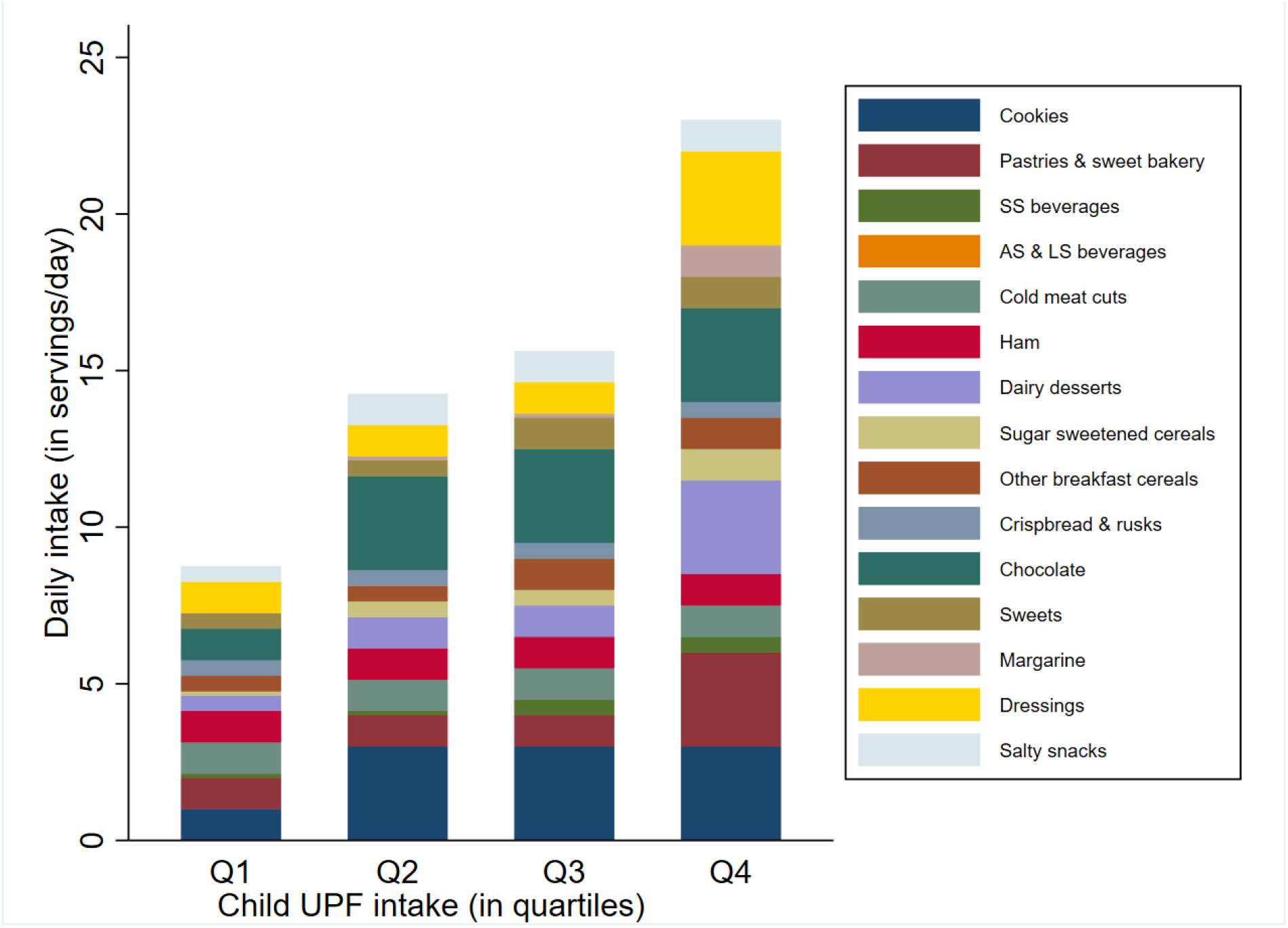
Median intakes (servings/day) of the foods and drinks identified as ultra-processed (UPF) by quartile of UPF intake. Quartiles of ultra-processed food intake (expressed as % of total daily food intake) were as follows: Q1, ≤18%; Q2, >18% but <23%; Q3, ≥23% but <29%; and Q4, ≥29%.

**Supplementary Figure 3.**
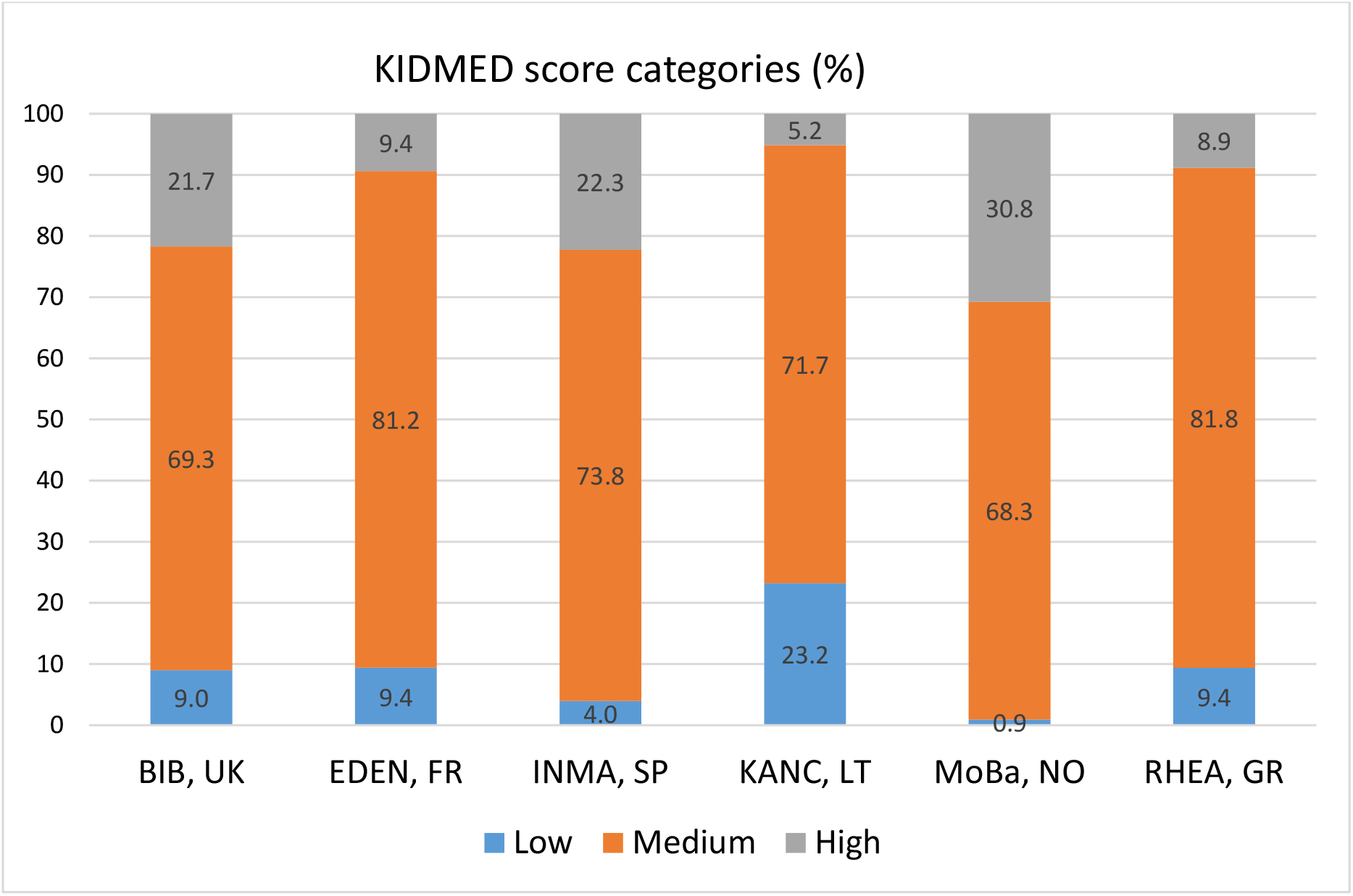
Levels of childhood adherence to the Mediterranean in each HELIX cohort. Adherence levels of Mediterranean diet were defined as follows: low, KIDMED score, <1; moderate, KIDMED score, 1-4; and high, >4.

**Supplementary Figure 4.**
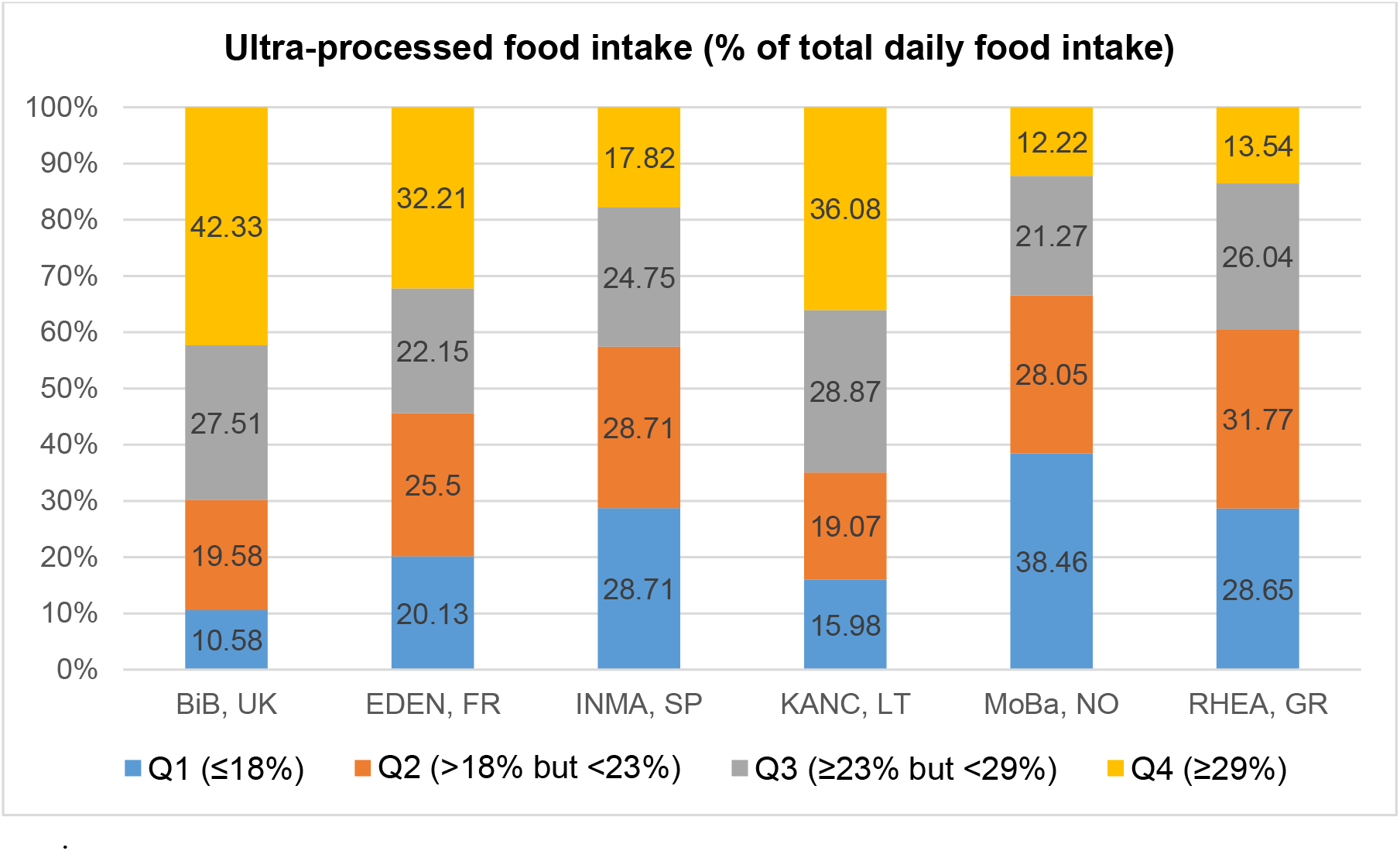
Levels of ultra-processed food consumption in each HELIX cohort. Levels of ultra-processed food intake (expressed as % of total daily food intake) are based on quartile cutoffs according to the overall intake distribution.

**Supplemental Figure 5.**
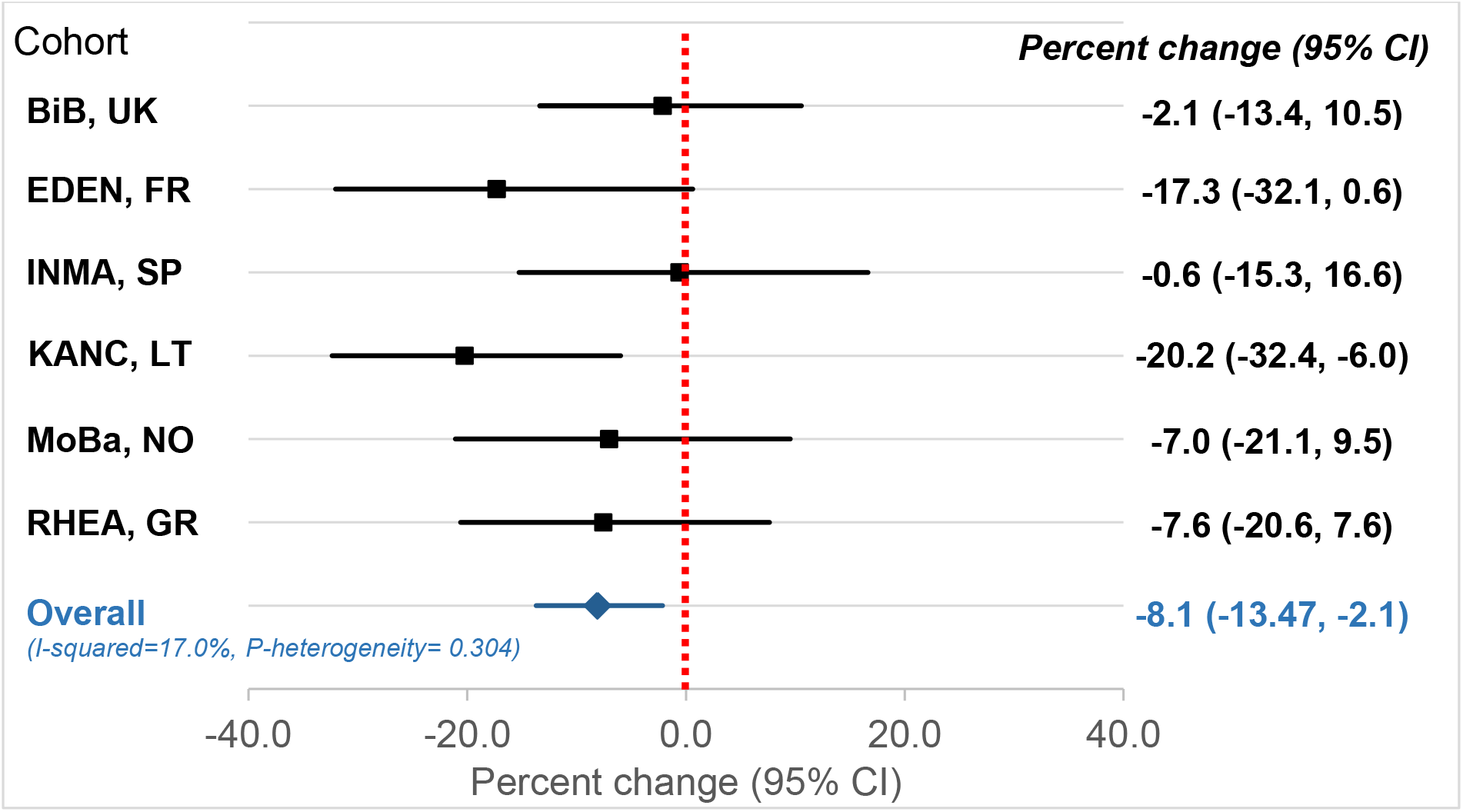
Cohort-specific associations of the KIDMED score with C-peptide in childhood. Beta coefficients (95% CIs) by cohort were obtained using linear regression models adjusted for maternal age, maternal education level, maternal pre-pregnancy BMI, family affluence status, child sex, child age, child BMI, child sedentary behavior, child ethnicity, and postprandial interval. Combined estimates were obtained by using a fixed-effects meta-analysis. Effect estimates were exponentiated and reported as percent change in C-peptide levels. Squares represent the cohort-specific effect estimates; diamond represents the combined estimate; and horizontal lines denote 95%CIs.

**Supplemental Figure 6.**
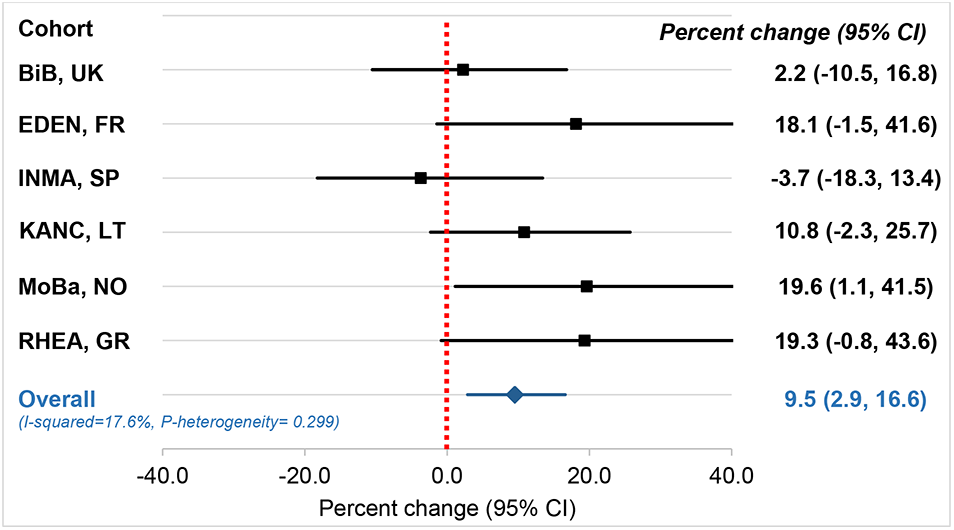
Cohort-specific associations of ultra-processed food intake with Cpeptide in childhood. Beta coefficients (95% CIs) by cohort were obtained using linear regression models adjusted for maternal age, maternal education level, maternal pre-pregnancy BMI, family affluence status, child sex, child age, child BMI, child sedentary behavior, child ethnicity, and postprandial interval. Combined estimates were obtained by using a fixed-effects meta-analysis. Effect estimates were exponentiated and reported as percent change in C-peptide levels. Squares represent the cohort-specific effect estimates; diamond represents the combined estimate; and horizontal lines denote 95%CIs.

## Notes

### Competing Interest Statement

The authors have declared no competing interest.

### Author Declarations

University of Southern California Institutional Review Board

